# A genetic map of human metabolism across the allele frequency spectrum

**DOI:** 10.1101/2025.01.30.25321073

**Authors:** Martijn Zoodsma, Carl Beuchel, Summaira Yasmeen, Leonhard Kohleick, Aakash Nepal, Mine Koprulu, Florian Kronenberg, Manuel Mayr, Alice Williamson, Maik Pietzner, C Claudia Langenberg

**Author notes:** Corresponding authors: Maik Pietzner, Claudia Langenberg. These authors contributed equally (Maik Pietzner, C Claudia Langenberg).

## Abstract

Genetic studies of human metabolism identified unknown disease processes and novel metabolic regulators, but have been limited in scale and allelic breadth. Here, we provide a data-driven map of the genetic regulation of circulating small molecules and lipoprotein characteristics (249 metabolic traits) measured using protein nuclear magnetic resonance spectroscopy (^1^H-NMR) across the allele frequency spectrum in ∼450.000 individuals. In trans-ancestry analyses, we identify 29,824 locus–metabolite associations mapping to 753 regions with effects largely consistent between men and women and major ancestral groups represented in UK Biobank. We develop a framework for classifying the observed extreme genetic pleiotropy, enabling identification of upstream ‘master’ regulators of lipid metabolism (’proportional pleiotropy’), such as *ANGPTL3.* We establish rare-to-common allelic series by integrating machine-learning guided effector gene assignments with rare exonic variant analyses providing high confidence gene assignments at >100 loci, including less established regulators of lipid metabolism like *SIDT2.* At 17 such loci we observed phenotypic heterogeneity among variants mapping to the same gene indicating differential metabolic roles of the altered gene product. We identify *VEGFA* as a potential modulator of HDL-mediated risk for coronary artery disease. Our results demonstrate how rare-to-common genetic variation combined with deep molecular profiling can identify unknown and inform on poorly understood regulators of human metabolism to guide prevention and treatment of diseases.

## Introduction

Our understanding of human metabolism is mostly based on dedicated hypothesis testing in experimental settings, informed by model organisms or observations in rare diseases patients. Only recently, high-throughput profiling of small molecules in large-scale studies has enabled systematic testing of genetic variation across the genome and provided an agnostic approach for the discovery of genes that encode key metabolic regulators^1–11^. These efforts have provided important new insights into how genetic variation shapes human chemical and metabolic individuality^1^ and have corroborated a large body of biochemical knowledge^1,2,10,12^.

The value of such genome-metabolome-wide association studies (mGWAS) extends beyond the mapping of biochemical pathways, sometimes demonstrating almost immediate clinical value. They provided examples how readily available supplementation strategies may prevent disease or delay onset in high risk individuals, such as serine for the rare eye disorder macular telangiectasia type 2^2^. Others further identified unknown variants affecting the absorption, distribution, metabolism, and excretion of exogenous compounds, most importantly drugs^1,13^, providing pathways to mitigate adverse drug effects. However, there are several challenges that currently limit the potential of mGWAS studies, in particular for causal inference. These include 1) the still rather small number of, at most, a dozen genetic variants linked to single molecules, 2) the inability to distinguish whether pleiotropic variants act on different molecules or pathways independently (horizontal pleiotropy), or whether they serve as ‘root causes’ of successive downstream changes (vertical pleiotropy), 3) the difficulty in distinguishing between locus-specific and metabolite abundance effects when colocalization at disease-risk loci is observed^1^, and 4) the challenge of confidently assigning effector genes at newly identified loci.

Here, we integrated rare (based on whole exome sequencing) and common genetic variation with measures of 249 metabolic phenotypes, including small molecules and detailed lipoprotein characteristics, among >450,000 UK Biobank participants representing three distinct ancestries. We demonstrate largely consistent genetic regulation across ancestries and sexes for almost 30,000 locus – metabolite associations and systematically categorise abundant genetic pleiotropy. By integrating machine-learning derived effector gene assignments with rare exonic variation, we identify previously unknown regulators of metabolism and observe heterogeneity in association profiles for variants mapping to the same gene. Finally, we demonstrate how systematic integration of statistical colocalization and Mendelian randomization can identify pathways with the potential to mitigate cardiovascular disease risk beyond current approaches focused primarily on LDL-cholesterol lowering.

## Results

We integrated genome-wide association studies (GWAS; population-specific minor allele frequency (MAF)≥0.5%) with rare exome-wide association studies (MAF≤0.5%) on plasma concentrations of 249 metabolite phenotypes, quantified using ^1^H-nuclear magnetic resonance spectroscopy (NMR). We included up to 450,000 UK Biobank (UKB) participants across three major ancestries (British White European – EUR (n=434,646); British African – BA (n=6,573); British Central South Asian – BSA (n=8,796); (**Supplementary Fig. 1**). The NMR measures provided a detailed readout of lipoprotein particles along a range of lipoprotein sizes containing 14 subclasses (i.e., extra-large very-low density (VLDL) to small high-density (HDL) lipoprotein particles), along with small molecules such as amino acids and ketone bodies quantified in molar concentration units (**Supplementary Table 1**).

### Common genetic variation underlying circulating metabolites

We identified 29,824 regional sentinel–NMR measure associations in trans-ancestral meta-analyses, representing 753 non-overlapping genomic regions (**Fig. 1a; Supplementary Table 2**). Nearly half of these regions (N=359, 47%) were associated with more than 10 NMR measures, demonstrating considerable pleiotropy cutting across metabolite classes for 350 regions. Characteristics of large HDL particles, such as concentration, particle size and (phospho)lipid, cholesterol, cholesteryl ester and triglyceride content, were associated with the largest number of regions (median: 166, IQR: 126-195), compared to median of 105 associated regions observed across all NMR measures (IQR: 68-142). Findings that considerably extended previous work^3^ and replicated parallel efforts using UK Biobank^9^ (**Supplementary Fig. 2**). Genes with well-characterised roles in human metabolism were significantly enriched among the closest genes to regional sentinels across different significance bins (adjusted p-values < 4.24 x 10^-9^; **Supplementary Fig. 3**). This suggests that ever-larger studies of often considered omnigenic traits, such as metabolites, still yield biological plausible findings and not merely non-specific upstream regulators.

**Figure 1:**
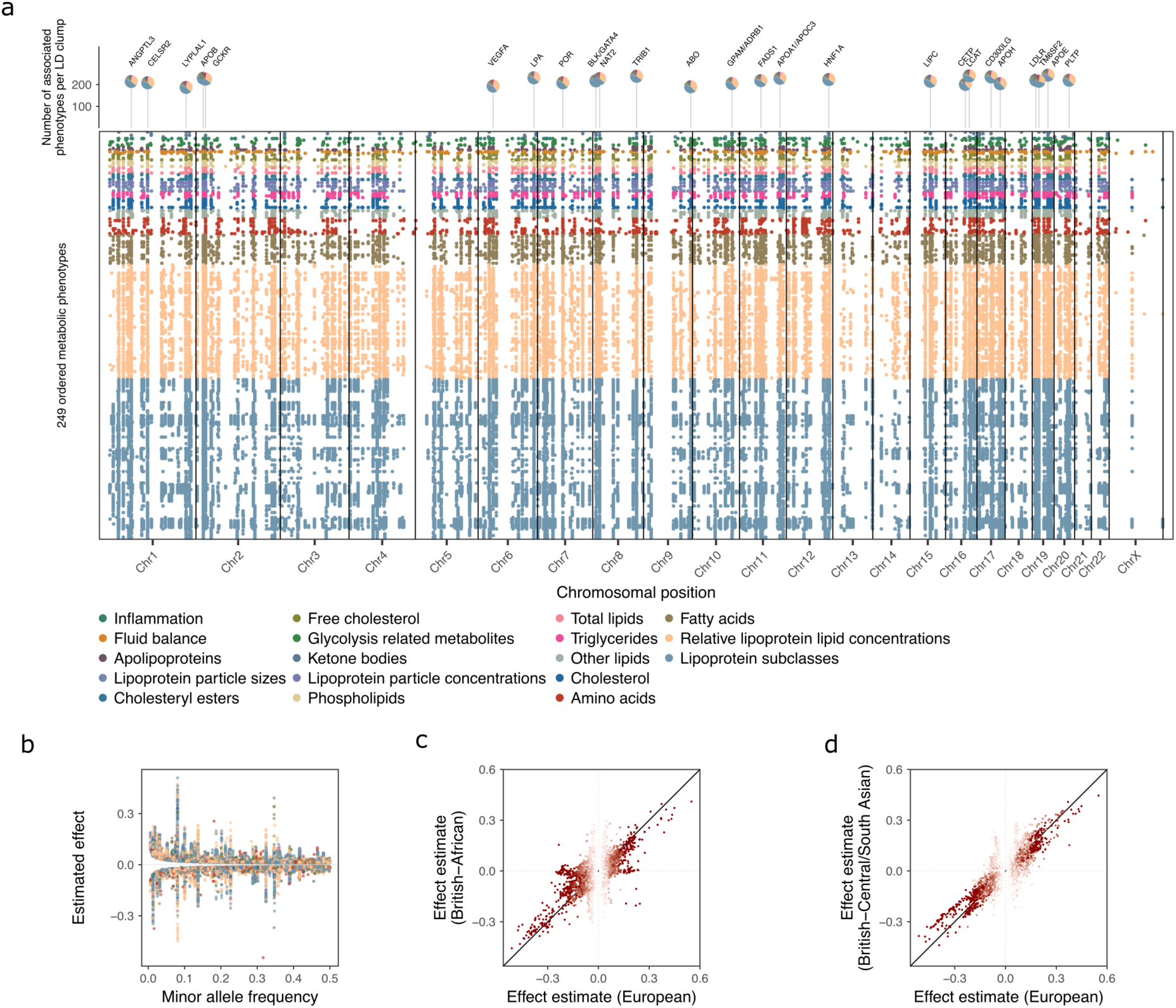
Common genetic regulation of circulating metabolites. **A)** Top-down Manhattan plot showing trans-ancestral sentinel variants for 249 metabolic phenotypes at a metabolome-adjusted genome-wide significance threshold of p < 2.0 x 10^-10^. Each row represents an NMR measure, coloured for biochemical class, chromosomal positions are shown on the x-axis. **B)** Weighted average allele frequency compared to estimated effect size for trans-ancestral sentinel variants. Points are coloured for biochemical classification. **C)** Comparison of effect sizes between White European samples (x-axis) and British-African samples (y-axis). We considered variants that were significant in either population. **D)** Similar to **C)** but comparing British-Central /South Asian samples. Dots are coloured according to their absolute Z-score in White European samples.

Almost all regional sentinel associations (n=29,410, 98.6%) showed little evidence of heterogeneity (p>10^-4^) across ancestries. To rule out possible artefacts that might have masked ancestral-specific effects (e.g., variant coverage and statistical power in the smaller ancestry groups), we repeated the GWAS within each ancestry separately. Consistent with the trans-ancestral meta-analysis, we observed high correlations of effect estimates for regional sentinels identified in the largest subgroup of White European participants when compared to those of African and Central South Asian ancestry (**Fig. 1c-d**; **Supplementary Table 3, Supplementary Fig. 4**). Although we note that the limited sample size did not permit comprehensive replication, our ancestry-specific analyses also revealed one locus not seen in European participants. The previously reported^14^ missense variant rs3211938 within *CD36* which is common in people of African ancestry (MAF_BA_= 0.12) but absent in European ancestry (MAF_EUR_= 0.0), was significantly associated (p-values < 1.49 x 10^-10^) with lower plasma concentrations of omega 3 fatty acids and 15 other NMR measures, including lipoprotein particle characteristics. This is in line with the role of CD36 as a fatty acid translocase, facilitating the recognition and uptake of long-chain fatty acids.

### Refinement of regional associations through multi-ancestry fine-mapping

We next employed a two-stage strategy to refine regional associations to a small number of candidate causal variants. Firstly, we implemented fine-mapping in the largest group of European-ancestry participants. We then further refined the subset of loci with at least suggestive evidence across ancestries (p<10^-4^) using trans-ancestral fine-mapping, leveraging the differential blocks of linkage disequilibrium (LD) despite vastly different sample sizes.

We first identified 3,007 statistically independent metabolite quantitative trait loci (mQTLs) associated with one or more NMR measure, representing a total of 43,322 credible set – NMR measurement pairs (**Supplementary Table 4**). This successfully defined 16,170 credible sets with a high-confidence variant (posterior inclusion probability (PIP) > 0.5). Among these were low-to-common frequent variants with functional consequences in metabolic genes, such as rs78734745 (MAF=0.8%; PIP=67.9%), a splice donor variant for *ME1*, associated with plasma citrate levels (beta=- 0.11; p-value<1.6×10^-21^). Lead fine-mapped mQTLs for a given NMR measure explained, on average, 6.9% (range: 0.57% - 13.42%) of the variance in plasma concentrations (**Supplementary Fig. 5**).

Secondly, we leveraged the different LD-block structure among participants of British African and British Central South Asian ancestry to further refine a total of 3,336 credible sets that still contained >1 variant and for which the locus had at least suggestive evidence for significance in either ancestry (P < 1.0 x 10^-4^). Trans-ethnic fine-mapping led to an increase in the number of credible sets containing high-confidence variants (Europeans: 997, multi-ancestral: 1,794) and decreased the median credible set size from 9 to 4 variants, while increasing the median posterior inclusion probability from 0.06 to 0.16 (**Supplementary Fig. 6**). This included 1,107 (33.7%) credible sets with two or fewer variants, and 1,518 (45%) credible sets that were reduced in size by more than half. We note, however, that most eligible European credible sets were already comparatively small (median 9 variants), but sometimes still spanned multiple genes.

For example, a signal associated with mono-unsaturated fatty acids (MUFA) concentrations at 17q21.2 contained 76 genetic variants spread across several genes covering a 1Mb window in the European-only discovery. The signal was fine-mapped to as few as 4 variants (two intergenic, one <50kb distance to the gene body) after incorporating evidence from other ancestries (**Supplementary Fig. 6**). Three of these four variants mapped to the *PTRF/CAVIN1* gene, which plays a crucial role in the formation of caveolae that are particularly abundant in adipocytes. Thus, *PTRF/CAVIN1* has been linked to generalized lipodystrophies^15^, providing a biologically plausible effector gene at this locus through trans-ancestral refinement of the credible set.

### Sex-differential effects at loci encoding metabolic genes

Many aspects of metabolism are known to vary by sex^16,17^, but only few genetic loci have been identified that may explain such differences^18,19^. While we observed highly correlated effect sizes across female and male participants (median R^2^: 0.98, range: 0.90 – 0.99), we also identified 360 putative sex-differential loci for 239 metabolic traits, representing 1,800 heterogenous associations in sex-stratified meta-analyses (heterogeneity p-value < 5 x 10^-8^, see **Methods**). To rule out that sex-differential effects could be explained by other factors that differ between the sexes, we performed additional analyses identifying that sex-differential effects at one-third of loci (n=625, 34.7%) were attenuated when controlling for factors such as body mass index, tobacco use, alcohol intake, and the use of lipid-lowering or diabetes medication (**Supplementary Fig. 7, Supplementary Table 5**). For loci unaffected by such additional factors, effect estimates were generally directionally concordant between the sexes but showed differences in magnitude (**Fig. 3a**). This is consistent with results previously observed for proteomics^20^ and suggests that the majority of significant sex interactions do not reflect sex-discordant effects. We observed pleiotropic sex-differential loci associated with 30 or more NMR measures near established lipoprotein genes (*APOE, APOC1, LPL*) but also less established genes (*SIDT2*), where sex was the most likely modifying factor. These finding may help to better understand sex-specific cut-offs in cardiovascular risk assessment in clinical guidelines to initiate treatment with lipid lowering medication^21^. We found *CPS1* on 2q34 to show the strongest sex differences, in line with previous reports^18^, with effect sizes for glycine being twice as large in females compared to males (rs1047891, beta females = 0.77, beta males = 0.34 s.d. units).

**Figure 3:**
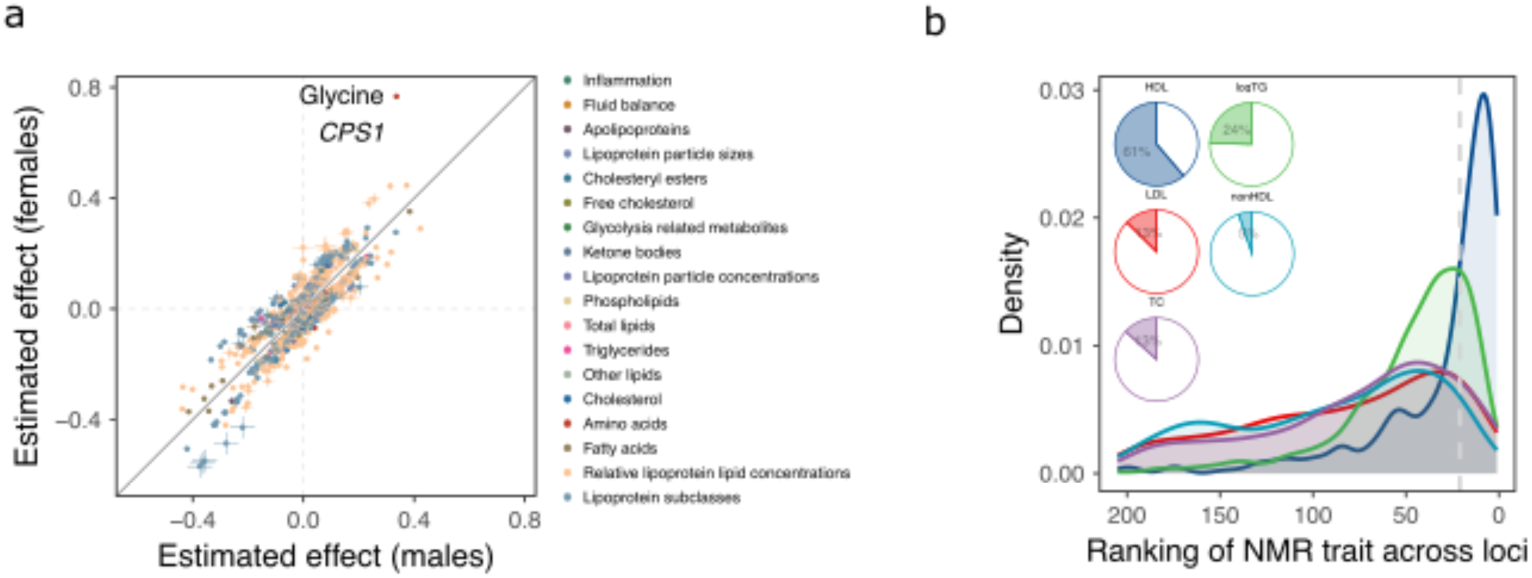
Putative sex-differential loci and reclassification of established lipid loci. **A)** Comparison of effect sizes of putatively sex-differential loci (defined as loci with heterogeneity p-value < 5 x 10^-8^ in a meta-analysis across the sexes). **B)** Rank distributions for each of the five matching NMR traits compared to the Lipids Genetics traits across genetic loci. Per locus – trait combination, 205 lipid-related NMR traits were ranked based on their absolute effect size and compared to the NMR trait that corresponds the Lipids Genetics consortium trait. Pie charts show the percentage of loci where the corresponding NMR trait is ranked among the top 10% of associated traits.

### Biological reclassification of established ‘lipid’ loci

To assess the value of metabogenomic studies involving lipoprotein profiling based on ^1^H-NMR spectroscopy over standard clinical markers, we systematically classified the NMR metabolome association profiles for 1,657 genetic variants reported for commonly measured clinical markers (LDL-cholesterol, HDL-cholesterol, total cholesterol and triglycerides) by the Global Lipids Genetics consortium (GLGC) in 1.6 million samples^22^. Around 25% of associated variants had the corresponding NMR measure among the top 10% of the most strongly associated NMR measures, with 22.5% of genetic variants showing significantly stronger associations with refined lipoprotein measures compared to their matching measure on the NMR platform, an observation most pronounced for non-HDL and LDL-cholesterol concentrations (**Fig. 3b**). While this indicated that relevant loci for lipoprotein metabolism can be discovered using readily available clinical measurements, it also demonstrates the necessity of refined lipoprotein profiles for better understanding the relevant biological pathways, including any inference about druggability or use for genetic causal inference methods. One such example was the *PNPLAP3* locus (tagged by rs3747207, associated with LDL-cholesterol by the GLGC; p = 2.3×10^-21^, beta = -0.014), where we observed no evidence of association with LDL-cholesterol (beta=-0.001, p = 0.49) but LDL particle size (beta=0.045, p-value = 1.04 x 10^-73^), and multiple characteristics of extra-large VLDL particles (**Supplementary Fig. 8**). The intronic rs3747207 variant is in strong LD (r^2^=0.98) with the well-known missense variant rs738409 (p.I148M) that has been demonstrated to confer hepatic lipid accumulation by altering ubiquniation of patatin-like phospholipase domain-containing protein 3 (PNPLA3) encoded by *PNPLA3*^23^. Our results provide human genetic support for a recently proposed role of *PNPLA3* in the secretion of large VLDL particles^24^. The association with LDL-cholesterol in massive scale studies likely being a distant downstream consequence.

### Machine-learning guided effector gene assignment

Assigning effector genes to genetic variants remains one of the most important bottlenecks for translating GWAS results into tangible insights. We assigned effector genes for almost three-quarters of European fine-mapped mQTLs (73.6%; n=2,213) with at least moderate confidence (candidate gene score ≥1.5, range 0 to 3), including about 28.2% with high-confidence assignments (score≥2; n=848), by training a machine learning model that integrates functional genomic resources with pathway information inspired by the ProGeM framework^25^ (**Supplementary Table 6**). For example, we prioritised the fatty acid elongase gene *ELOVLC* for 16 different NMR measures (tagged by rs3813829), including the fraction of cholesterol and other fatty acids on very small VLDL and very large HDL particles in addition to the fraction of saturated fatty acids. The gene product, ELOVL fatty acid elongase 6, catalyses the rate-limiting step in long-chain fatty acid elongation, which are subsequently incorporated into lipoprotein particles. We also prioritized genes with upstream roles in metabolism, including a locus on 17q25.3 where we prioritized cytohesin-1 (*CYTH1)* as the candidate causal gene for five independent, genetic variants linked to 11 distinct NMR measures mostly comprising characteristics of VLDL particles. *CYTH1*, previously associated with type 2 diabetes^26^, promotes activation of ADP-ribosylation factors (ARF)1, ARF5 and ARF6, regulators of lipid vesicle transport, membrane lipid composition and modification^27^, demonstrating a relevant but indirect link to lipoprotein metabolism.

We observed considerable overlap of machine-learning guided effector gene predictions (top three genes) with those reported based on manually curated biological plausibility (191 out of 283 loci)^3^ or based on colocalization with protein quantitative trait loci that have not been used to train the algorithm^28^ (81 out of 143; **Supplementary Table 6**). While missing overlap indicates room for improvement, 24 high-confidence assigments did strongly disagree with either external source (gene score >2 but no match among pQTL prioritised or manually curated ones). This included a locus on chromosome 19q13.11 tagged by rs62102718 for which we prioritised *PEPD* with high-confidence (score=2.42) as opposed to *CEBPA*^3^. PEPD encodes peptidase D, highly relevant for collagen turnover, that has been shown to promote adipose tissue fibrosis in mouse knock-out models and promoting insulin resistance^29^. Insulin resistance, in turn, being a very plausible explanation for the pleiotropic effect of the variant on diverse lipoprotein characteristics (n=31).

### Tissue distribution of effector genes

We next tested tissue-specific expression patterns of the identified effector genes to better understand organ sites contributing to (lipoprotein) metabolism. Strong clustering was observed at both the tissue and metabolite levels, reflecting both known and less established organ contributions (**Supplementary Fig. 9a, Supplementary Table 7**). Genes characteristic of the liver, adipose tissue, adrenal gland, but also female breast tissue (likely reflecting its high adipose tissue content) were significantly enriched among effector gene sets across the metabolic measures captured by NMR. This included significant enrichment of all amino acids in liver tissue (e.g., phenylalanine: odds ratio (OR): 14.8, p<1.3 x 10^-8^, histidine: OR 7.9, p<2.9 x 10^-11^) but also for skeletal muscle in alanine metabolism (OR:3.82; p-value<7.9×10^-9^). Similar enrichments were observed when using the closest gene instead of our annotated effector genes for mQTLs (**Supplementary Fig. 9b**).

### Modes of metabolic and systemic pleiotropy

Pleiotropy is a widespread but poorly understood phenomenon and we developed a framework to characterise four different modes of metabolic pleiotropy for all fine-mapped mQTLs (**Fig. 4a-d**; **Supplementary Fig. 10 and Table 6**; **see Methods**). About half of the pleiotropic mQTLs (n=880; ≥2 NMR measures) showed evidence for two different modes of vertical pleiotropy. Firstly, within confined pathways (n=218; ‘pathway pleiotropy’) or, secondly, as a function of the correlation with the ‘lead’ NMR measure (n=662; ‘proportional pleiotropy’; **Fig. 4a**). For example, rs76594121 tagged an mQTL at 3q21.3 associating with different characteristics of large HDL particles, for which we prioritized *ACAD9* as the most likely candidate gene (**Fig. 4a**). The gene product of *ACAD9*, acyl-CoA dehydrogenase family member 9, is part of complex I of the respiratory chain that catalyses the oxidation of fatty acids with a high affinity for long chain fatty acids that are, amongst others, carried by HDL particles. A prototype example for ‘proportional pleiotropy’ was an mQTL tagged by rs624698 for which we prioritized *ANGPTL3* as the effector gene (**Fig. 4b**). Angiopoietin-like 3, encoded by *ANGPTL3*, inhibits lipoprotein lipase activity but also endothelial lipase, resulting in increased triglycerides, HDL-cholesterol, and phospholipid concentrations, consistent with HDL-particle characteristics being the most strongly associated NMR measures (p<1.0×10^-546^). Other associations being downstream effects on lipoprotein metabolism rather than acting on independent pathways (**Fig. 4b**), considerably expanding previous genetic observations^30^.

**Figure 4:**
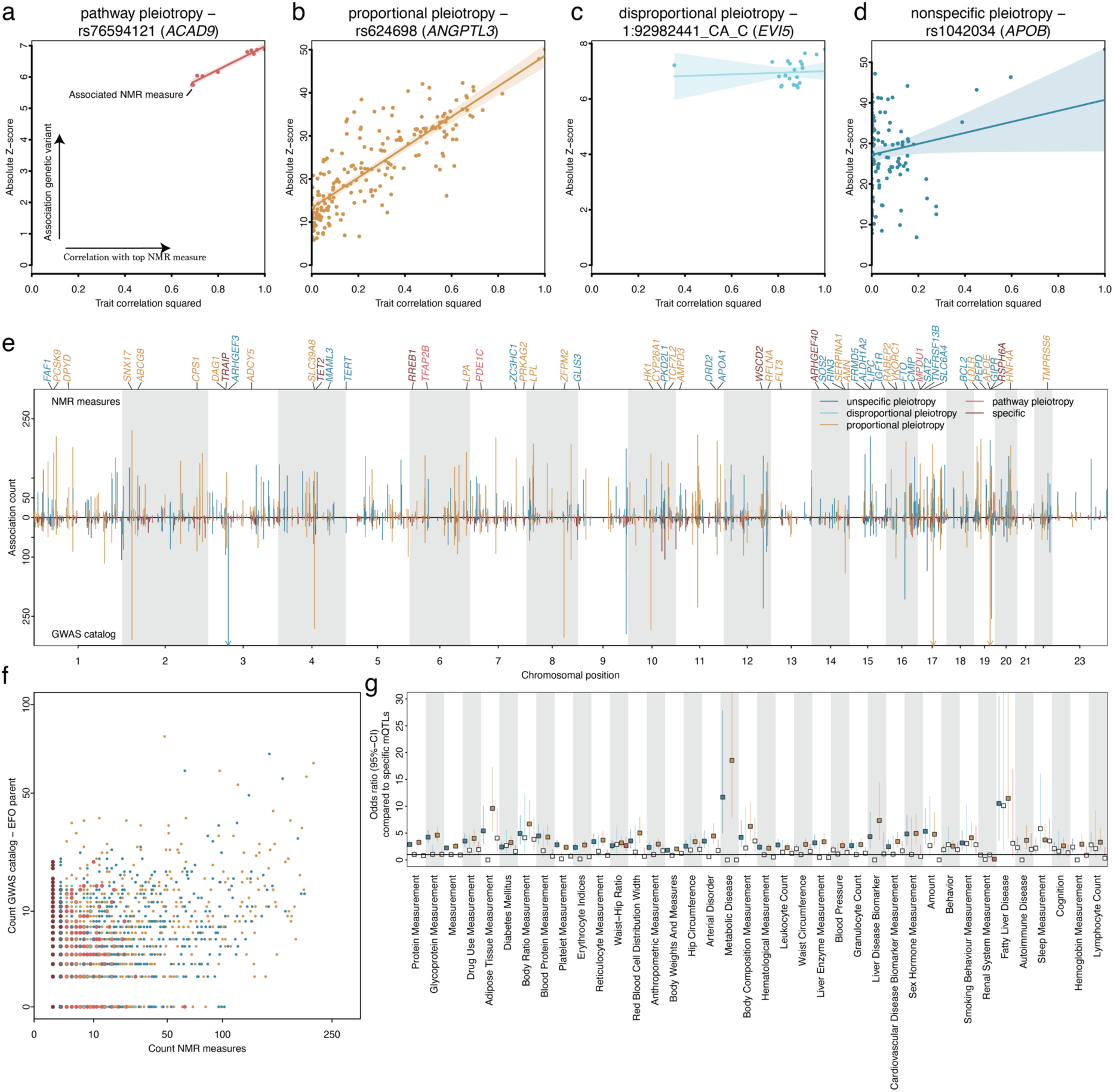
Modes of pleiotropy. **a-d)** Exemplary scatterplots opposing the squared trait correlation of the lead NMR measure for the listed variant against the absolute Z-score from linear regression models for all associated NMR measures. The colours indicate different modes of pleiotropy and correspond to the legend in e). For each plot, a linear regression fit with 95%-confidence interval is given. **e)** Number of associated NMR measures for each of 3007 mQTL groups opposed to associations reported in the GWAS catalog after pruning the GWAS catalog for metabolic phenotypes (see Methods). Colouring is according to modes of pleiotropy. **f)** Scatterplot opposing the number of associated NMR measures (x-axis) of each mQTL group with the number of reported EFO parent categories in the GWAS catalog. **g)** Odds ratios and 95%-confidence intervals from logistic regression models testing whether EFO categories (x-axis) are more frequently reported for pleiotropic mQTL groups compared to specific ones. Darker colours indicated estimates passing corrected statistical significance.

The remaining half of pleiotropic mQTLs showed evidence for two modes of horizontal pleiotropy: those with evidence for ‘disproportional pleiotropy’ (n=68) and a larger group with evidence for ‘nonspecific pleiotropy’ (n=720). For example, a small deletion on chromosome 1 (chr1:92982441:CA>C) was associated with a highly correlated cluster of NMR measures, including characteristics of IDL, LDL, and VLDL particles (**Fig. 4c**), but for which we detected no correlation of association strengths according to the lead NMR measure, the concentration of esterified cholesterol in medium-sized VLDL particles (p<6.8×10^-14^). We prioritized *EVI5* as the most likely candidate gene, supported by previous studies on rare functional variants^31^. The gene product of *EVI5*, ecotropic viral integration site 5, has no apparent link to (lipoprotein) metabolism in line with most of the gene assignments for mQTLs with a similar nonspecific pleiotropy pattern. An example of ‘nonspecific pleiotropy’ was the *APOB* missense variant rs676210 (p.Pro2739Leu) associated with 126 NMR measures across the entire lipoprotein density range, but also creatinine and glycoprotein acetyl concentrations (**Fig. 4d**). The differential effects of the same genetic variation on distinct lipoprotein subgroups aligns with changes in lipid profiles seen with mipomersen, an antisense oligonucleotide against *APOB*, that demonstrated reductions in LDL-cholesterol but also subsequent increases in the triglyceride content of VLDL particles as hepatic adaption occurs^32^.

Modes of molecular pleiotropy only partially translated into pleiotropy across the entire breath of phenotypes and diseases studied genetically (**Fig. 4e**). We observed a two-fold enrichment of ‘proportional pleiotropic’ (OR: 2.11; p<2.0×10^-14^) and to a lesser extend an enrichment of ‘nonspecific pleiotropic’ (OR: 1.52; p<1.1×10^-5^) variants among variants reported in the GWAS catalog for ≥5 non-metabolomic trait categories (**see Methods**). In contrast, the set of pleiotropic GWAS catalog variants was significantly depleted for ‘specific’ mQTLs (odds ratio: 0.42; p<1.6×10^-21^). Some phenotypically specific variants thereby provided clues to understand non-specific molecular pleiotropy (**Fig. 4f**). For example, rs8101064, an intronic variant in *INSR,* encoding the insulin receptor, has been reported for type 2 diabetes among East Asians^33^ and was associated with 40 NMR measures in a nonspecific manner, likely reflecting the broad effects of insulin resistance on whole body lipid metabolism. Systemic mechanisms explaining effects of ‘proportional’ and ‘nonspecific’ pleiotropic mQTLs were further evidenced by a more than 20-fold significant enrichment of associated trait categories such as ‘metabolic disease’, ‘fatty liver disease’, and ‘arterial disorders’ (**Fig. 4g**).

### Convergence of common and rare genetic variation shaping metabolism

Previous investigations focussed on either large-scale common variant^1–4,8^ or comparatively small-scale rare exonic variant discovery efforts^7,34^, but these approaches have not been able to integrate such information at scale to establish allelic series that confidently link genes to metabolites, including previously unknown regulators of metabolism. We identified rare variation (MAF≤0.05%) in a total of 209 genes to be significantly (p< 1.1 x 10^-8^) linked to one or more of 249 NMR measures combining ultra-rare gene burden analysis (3,709 significant associations; **Supplementary Table 8**) and rare exonic variant analysis (4,131 significant associations; **Supplementary Table G**). Effect sizes were significantly larger compared to more frequent variant effects (**Fig. 5a**). For example, people carrying rare predicted loss-of-function variants in *SLC13A5* had more than 1.4 s.d. units higher plasma citrate concentrations per copy of the possibly damaging allele (beta:1.41; p-value<2.6×10^-20^).

**Figure 5.**
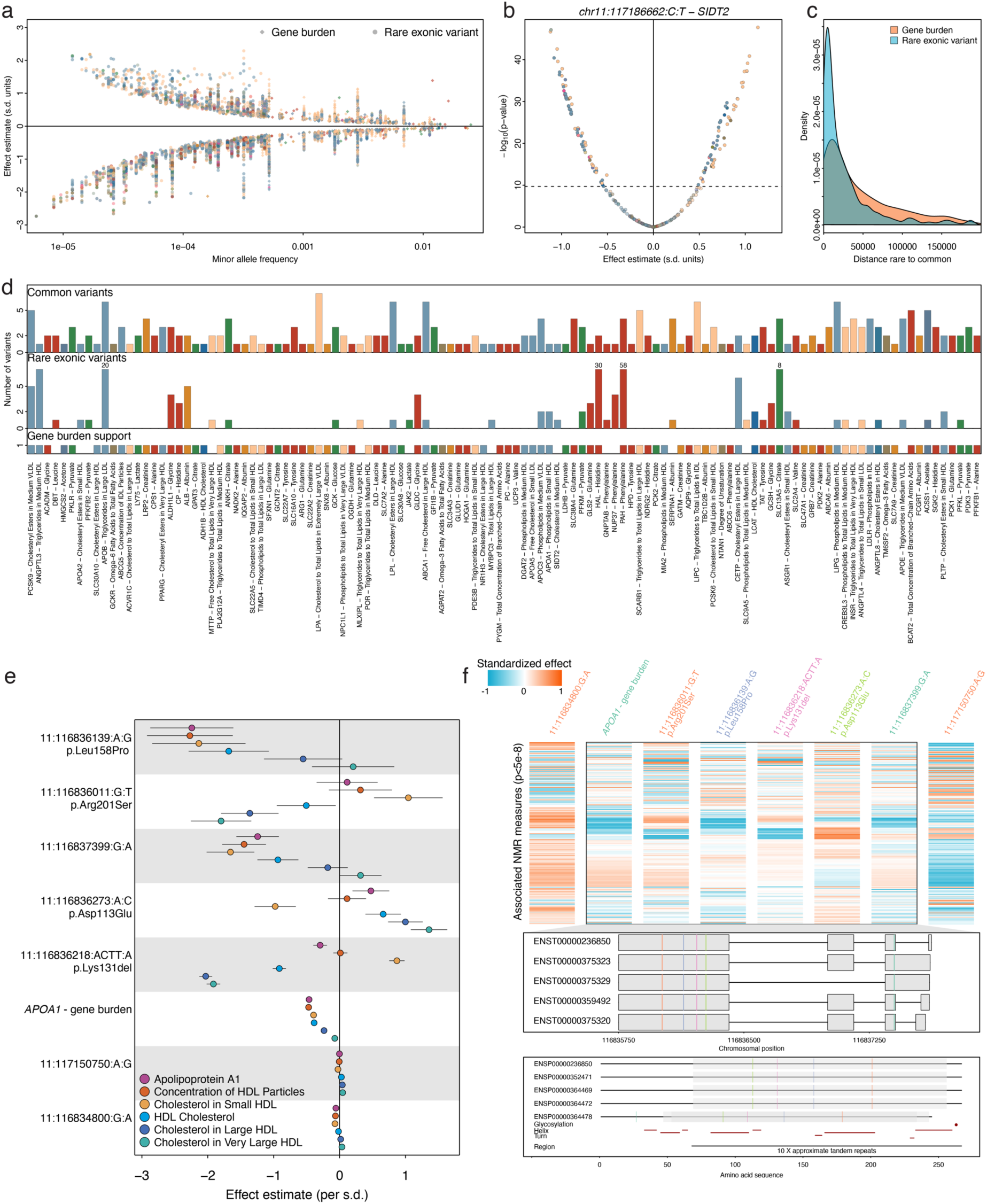
Rare coding variation associated with NMR measures and convergence with common variant associations. **a)** Effect estimates against minor allele frequency (MAF) of significantly associated gene burden (diamonds; p<1.2×10^-8^ and rare exonic variants (MAF<0.05%; circles; p< 2.0 x 10^-10^). **b)** Effect estimates and -log10(p-values) for associations of the rare intronic variant chr11:117186662:C>T within *SIDT2* across all 249 NMR measures. The dotted horizontal line indicates the multiple testing threshold (p<2.0×10^-10^). **c)** Genomic distance between gene burden (blue) or rare exonic variants (orange) towards the next common credible set variant. **d)** Evidence for allelic series based on i) gene burden analysis (bottom panel), ii) rare exonic variants (middle panel), and iii) common variants with prioritized effector gene matching to the evidence from exonic analysis. For each gene, only the NMR measure most significantly associated with the strongest common variant is shown in case multiple NMR measures were associated. Some bars for the number of associated rare exonic variants have been capped to fit into plotting margin but the number is given in the plot. **e)** Effect estimates (dots) and 95%-confidence intervals for 7 variants mapping to *APOA1* as well as a cumulative burden of high-confidence pLOF variants within *APOA1* and bespoke circulating measures of ApoA1 and HDL particles (colour gradient). **f)** The top displays a heatmap of standardized effect estimates (per variant) across 87 NMR measures for each associated variant and a cumulative burden within *APOA1*. Variants mapping into the region encoding the protein are surrounded by a rectangle. Variant effects have been aligned to the minor allele. The middle panel maps the corresponding variants to their respective transcripts encoding different forms of *APOA1*, while the lowest panels maps missense variants onto the amino acid sequence of the protein. Variant names coloured similarly had highly correlated association profiles.

We also observed considerable pleiotropy, including 47 genes associated with 20 or more NMR measures. Many of these genes have well-known roles in metabolism or small molecule transport, such as half (n=23/51) of the genes being involved in (peripheral) cholesterol metabolism (**Supplementary Fig. 11**). On the other hand, rare pleiotropic variants with large effect sizes (MAF < 0.02% and beta > 0.6 s.d. units) pointed towards less-established regulators of metabolism including *SIDT2* (chr11:117186662:C>T, n=124 carriers), *JAK2* (chr9:5073770:G>T, n=73 carriers) or *CEP164* (chr11:117356670:C>G, n=49 carriers). Experimental work already suggested a role for the gene product of *SIDT2* (SID1 transmembrane family member 2) in hepatic lipid metabolism and apolipoprotein A1 (ApoA1) secretion, the main protein component of HDL particles which constituted the majority of associated NMR measures (**Fig. 5b**)^35,36^. In contrast, associations with *JAK2* variants indicate a link to clonal haematopoiesis of indeterminate potential (CHIP)^37^ with uncertain causality.

We observed strong overlap between our gene burden and common variant findings, with 85.4% of rare variant (n=3528) and 75.5% of gene burden (n=2802) associations being no more than 100kb away from the nearest statistically independent lead credible set variant (**Fig. 5c**). In contrast, most common variant findings (92.3%) were not within 500kb of matching rare variant/burden evidence. Notably, 12.1% of gene burden results were more than 1Mb away from the next common credible set variant for the respective NMR measure, aligning with recent observations that both approaches partly prioritise different genes^38^.

At 116 genes (55.5%), rare variant and/or burden evidence overlapped with effector gene predictions for closeby common credible set variants (≤200kb) for one or more associated NMR measure (**Fig. 5d**), providing independent support for allelic series (**Fig. 5d; Supplementary Table 10**). For example, we identified an allelic series composed of 7 rare loss-of-function (LoF), 1 gain-of-function (GoF), and 4 common variants for serum citrate levels at *SLC13A5* encoding a sodium-dependent citrate co-transporter. Another allelic series at*ANKH* comprised four common variants (rs185448606 – MAF=1.3%; rs17250977 – MAF=4.0%; rs826351 – MAF=44.3%; rs2921604 – MAF=45.9%) and a rare missense variant chr5:14745916:T>C (MAF=0.0069%) being also associated with lower serum concentrations of citrate (beta=-2.18 s.d. units, p<5.2×10^-11^) (**Fig. 5d**). *ANKH* encodes for a multipass transporter, recently shown to transport citrate^39^, with an important role in bone health^39^.

We observed evidence that genetic variants even within an allelic series had differential metabolic consequences, covering a total of 17 genes associated with ≥10 NMR measures (**Supplementary Table 10**). The most outstanding example included 7 variants (5 rare; 2 common) and a cumulative burden of rare predicted LoF variants mapping to *APOA1*. They distinctively associated with one or more of 87 NMR measures, most strongly with diverse characteristics of HDL particles of which the gene product, Apolipoprotein A1 (ApoA1), is the major component (**Fig. 5e-f**). This included four rare missense variants (MAF≤0.03%) encoded in exon 4 that each had partly differential effects on the number, size, and cholesterol content of HDL particles (**Fig. 5e**). Only one of which (p.Leu158Pro) primarly associated with serum ApoA1 concentrations and HDL particle number, micking the association with the cumulative burden of high-confidence predicted LoF variants in *APOA1*, suggesting a potentially dysfunctional protein that lacks interaction with lecithin cholesterol acyl transferase to facilitate cholesterol uptake^40^. In contrast, p.Lys131del and p.Arg201Ser seemed to rather predispose to a shift in cholesterol content from large towards small HDL particles, a pattern opposed by p.Asp113Glu (**Fig. 5e**). An observation consistent with amyloid formation by ApoA1 that has been observed in early case reports of p.Lys131del (historically the ApoA-I_Helsinki_^41^) in which HDL-cholesterol or ApoA1 concentrations are only mildly changed but aggregation of misfolded ApoA1 protein can confer organ damage later in life^42^). Since p.Asp113Glu and p.Arg201Ser have not yet been identified to cause amyloidosis, we cannot rule out the possibility that each variant maps to distinctive parts of ApoA1 with subsequently different consequences on function and/or stability (**Supplementary Fig. 12**).

### Phenotypic consequences of rare variation in metabolic genes

Rare inborn errors of metabolism are among the few disorders screened for at birth by most healthcare systems globally, as early intervention – such as appropriate substitution or dietary regimens – can prevent developmental issues and diseases later in life. We observed a more than 3-fold enrichment of genes previously linked to Mendelian diseases^43^ (’OMIM genes’) among those associated with NMR measures in gene burden and rare exonic variant analyses (odds ratio: 3.30; p-value<6.5×10^-17^; **Supplementary Table 11**), in line with results reported from previous mGWAS^1,2,7,8^. For 15 out of 106 genes, we found evidence of significantly associated disease risk (p<7.5×10^-7^), largely replicating signs and symptoms of corresponding rare disorders (**Supplementary Table 12**). Associations with NMR measures thereby represented different modes of action. For cardiovascular diseases, most prominently familial hypercholesterolemia (e.g., via *APOB*), they likely acted as mediators, whereas associations converging on *PKD1* for cystic kidney disease likely indicated disease consequences. We further observed less understood pleiotropic roles of OMIM genes. For example, rare predicted loss-of-function variants within *SMAD6* are known to cause, amongst others, malformations of bones, e.g., Craniosynostosis 7, characterised by malformations of the skull and subsequent brain damage, and we observed a strongly increased risk for other disorders of the cervical region (OR:28.8; 95%-CI: 10.3 – 80.5; p-value<1.4×10^-10^), as well as significantly smaller VLDL particles (beta:-0.13; 95%-CI: -0.16 - -0.09; p-value< 1.5×10^-9^) among rare variant carriers in UKB. The gene product, SMAD Family Member 6, suppresses TGF-beta signalling, which has known effects on bone morphogenetic proteins^44^. Independent evidence suggests that *SMADC* downregulation reduces the expression of core genes involved in lipoprotein metabolism, such as *LDLR* ^45^, that may explain the disease-unrelated association.

When we tested more generally whether a rare variant burden in metabolic genes was associated with disease susceptibility, we observed a significant enrichment among susceptibility genes for endocrine and metabolic disorders, such as type 2 diabetes and different lipidemias but not among other disease categories (**Supplementary** Fig. 13).

### Risk mitigation of atherosclerotic cardiovascular disease beyond LDL-cholesterol

The success of LDL-cholesterol-lowering drugs for the prevention of atherosclerotic cardiovascular disease (ACVD) can be effectively recapitulated by genetic evidence. Genetic predisposition to high LDL-cholesterol is strongly associated with an increased risk of ACVD (‘level effect’), and genetic variations that mimic potent drug targets, such as at *PCSK9*, show strong evidence of shared effects on both LDL-cholesterol and ACVD (’locus effect’)^46^. To identify potential pathways to mitigate the residual risk not addressed by lowering of LDL-cholesterol^47^, we systematically integrated outcome data across 25 CVD phenotypes^48–62^, including non-atherosclerotic diseases to test specificity of disease associations, with NMR phenotypes and assessed the convergence of locus and level effects (**Supplementary Table 13**).

We identified significant evidence (false-discovery rate (FDR)<5%) for 1,146 ‘level effects’ across 218 NMR measures with one or more of 22 CVD phenotypes using pleiotropy curated genetic instuments in Mendelian randomization (**Fig. 6a; Supplementary Table 14**). Independently, we observed evidence for 5,527 ‘locus effects’, establishing a shared genetic architecture (posterior probability (PP)>80%) between 87 mQTL associated with 247 NMR measures and 17 CVD phenotypes (**Fig. 6b; Supplementary Table 15**). For a total of 46 NMR measure – CVD combinations we found converging evidence for level- and locus-effects, including 23 not associated in our study with parameter of LDL-metabolism (see Methods; **Fig. 6b**), providing potential alternatives for addressing residual risk (**Supplementary Table 16**).

**Figure 6.**
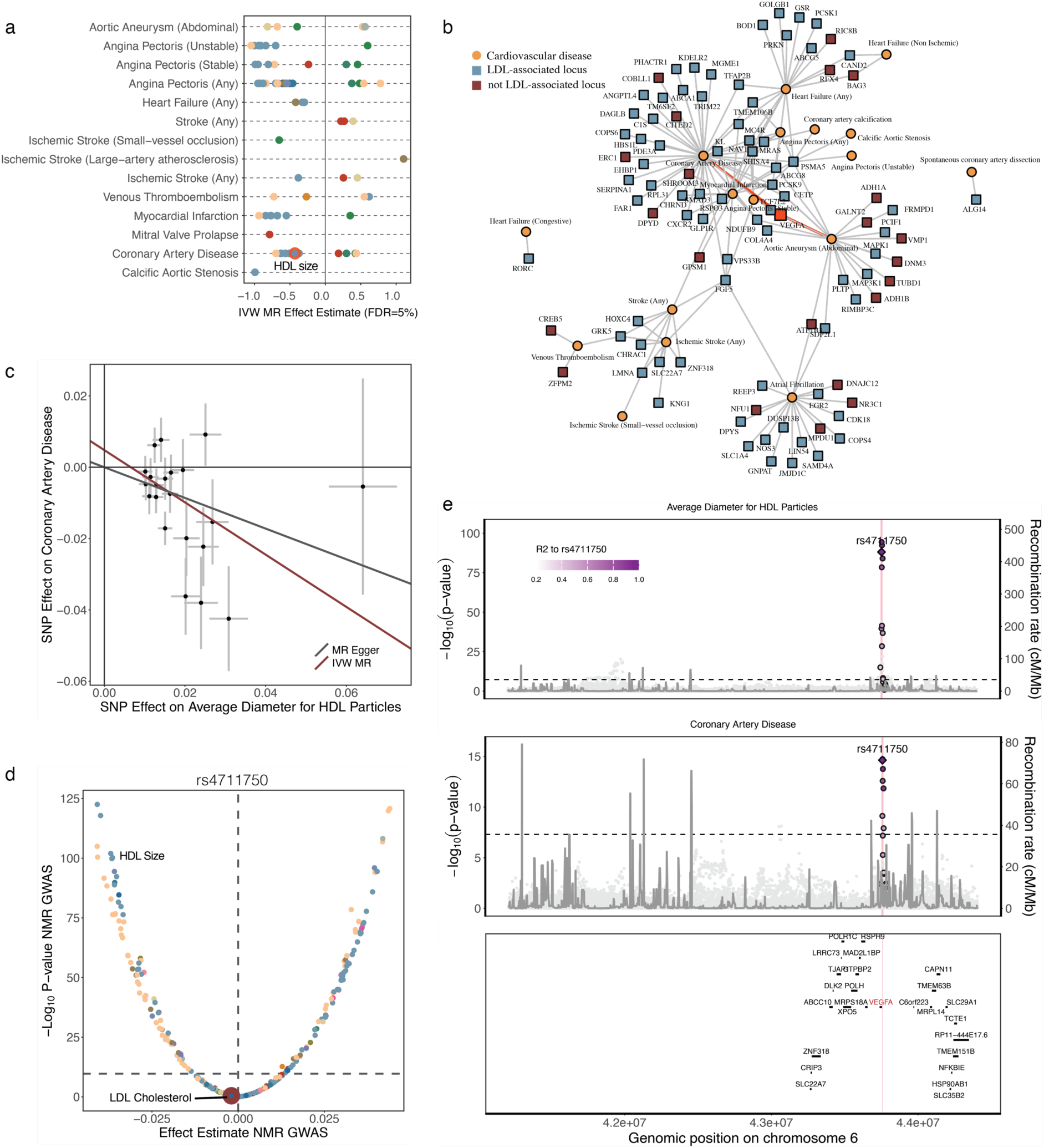
Genetic prioritisation to target residual cardiovascular risk. **a)** Summary of two-sample Mendelian randomization analysis testing for putatively causal effects of NMR measures on the risk on diverse cardiovascular diseases (CVD). Shown are effect estimates for NMR – disease pairs passing multiple testing. Metabolites are coloured according to the scheme from Figure 1. **b)** Locus – disease network highlighting loci for which at least one NMR measure showed evidence of colocalization with one or more CVDs (PP≥80%). Only loci without evidence for unspecific pleiotropy are depicted. Loci were annotated with the most likely causal gene. Loci coloured in blue showed evidence for being associated with LDL-Cholesterol whereas red did not. **c)** Dose-response plot for SNPs associated with HDL particle size (after filtering for pleiotropic SNPs) against the risk for coronary artery disease. Effect estimates (dots) and 95%-Cis are given and MR-regression lines added. **d)** Effect of rs4711750 across the NMR metabolome. **e)** Locuszoom plot centred around *VEGFA* demonstrating colocalization for the genetic signal for HDL particle size and coronary artery disease.

For example, we observed robust evidence that, among other measures related to HDL size and composition, genetic susceptibility to larger HDL particle size was associated with a 35% reduced risk of coronary artery disease (CAD; odds ratio=0.65; 95%-CI: 0.50 – 0.83; p_adj_<0.007, **Fig. 6c**) along with robust evidence of a shared and directionally concordant genetic signal tagged by rs4711750 at the *VEGFA* locus (PP = 99%, **Fig. 6e**). The locus has previously been implicated in CAD risk^48^, and our results now suggest that one likely pathway to modulate CAD risk might be via HDL particle size or characteristics of large HDL particles not captured by HDL-cholesterol. Vascular endothelial growth factor A (VEGFA), encoded at *VEGFA*, is primarily known for its role in angiogenesis^63^, but it has also been described as a regulatory factor of transendothelial transport of esterified cholesterol from HDL but not LDL particles via activation of scavenger receptor BI (SR-BI) during reverse cholesterol transport^64^. Inhibition of VEGFA is a major pharmaceutical target to suppress vascularisation of malignant tumours^63^, and agents targeting VEGF signalling are well-known for adverse cardiovascular effects^65^, suggesting that activation of VEGFA, rather than inhibition, might be necessary to potentially reduce CAD risk. Our observations contribute to a growing body of evidence that more tailored approaches - rather than increasing HDL cholesterol content – will likely be needed for potential cardiovascular benefits, given the discouraging trials for most agents increasing HDL-cholesterol^66^. We note, however, that HDL-particle size might still only be a ‘measurable’ surrogate, rather than being the true underlying mechanism. For example, inhibition of reverse cholesterol transport via dysfunctional SR-BI increased HDL particle size as well as CAD risk^67^.

### Disease-wide Mendelian randomization screen for non-lipoprotein measures

Having established categories of pleiotropy for mQTLs beyond simple association counting, we finally aimed to demonstrate its application in a disease-wide screen using 1394 disease outcomes from the FinnGen study^68^ (relase 11) for non-lipoprotein measures. We observed a strong decline, 29 to 13 metabolite – disease association with significant evidence (adjusted p-value < 0.05) from two-sample MR (‘level effect’) once subsetting to metabolite-specific instruments, indicating false-positive results due to pleiotropy (**Supplementary Table 17**).

We observed evidence for convergence of locus and level convergence for a risk-increasing effect of genetically predicted plasma glycoprotein acetyl concentrations on type 2 diabetes risk (odds ratio per 1 s.d. increase: 1.67; p-value<3.9×10^-7^). The association persisted even after additional exclusion of variants with evidence for pleiotropy in the GWAS catalog (odds ratio: 1.69; p-value<9.1×10^-5^). Notably, ‘locus’ convergence was based on the consistent effect of the rare loss-of-function variant chr20:44413714:C>T (MAF = 0.02%) within *HNF4A* on plasma glycoprotein acetyl concentrations (beta: 0.60; p-value<8.3×10^-15^) and the cumulative effect of ultra-rare loss-of-function variants on type 2 diabetes risk (odds ratio: 2.68; p-value: 6.5×10^-10^). However, we note that plasma glycoprotein acetyl concentrations proxy a complex chronic inflammatory state^69^ warrants further follow-up analysis to establish mechanistic links to type 2 diabetes. In contrast, previously reported associations between genetically predicted levels of branched-chain amino acids and type 2 diabetes reached at best nominal significance with a smaller effect size than previously estimated^70^ (e.g., plasma leucine concentrations: odds ratio per s.d. unit: 1.19; p-value<0.02).

## Discussion

The genetic basis of circulating metabolites provides insights into the complexion of human metabolomic regulation and its subsequent influence on health and disease. By integrating common and rare genetic variation with circulating metabolite concentrations in 450,000 individuals from three different ancestries, we provide here a data-driven map of the circulating metabolome across the allele frequency spectrum. This map identifies previously unrecognized modulators of metabolism with potential health implications.

By combining ML-guided common variant-to-gene annotation with rare exonic variation, we provided high-confidence effector gene assignments at >100 loci, including some with less establised roles in (lipoprotein) metabolism, such as *SIDT2*. These findings present compelling candidates for further functional studies, with a strong incentive that they are likely relevant to human biology, in contrast to species differences frequently encountered in animal models^71,72^. Large-scale studies similar to ours, but with a broader coverage of the plasma metabolome, will likely uncover many more genes with yet undefined roles in metabolism, complementing hypothesis-driven research in experimental models.

After more than two decades of GWAS, it has become clear that pleiotropic effects of genetic variants are ubiquitous (see, e.g., ^73^). Little distinction has been possible beyond the generic concepts of ‘vertical’ and ‘horizontal’ pleiotropy or measures of simple counting. We refine these concepts by observing variants associated with dozens of NMR measures but consistent with the concept of effects diluting/propagating along pathways (’proportional pleiotropy’). Conversely, we also observe variants associated with comparatively few NMR measures in an inconsistent pattern (’disproportional pleiotropy’) suggesting distinct effects on otherwise highly correlated traits. Our data-driven approach thereby augments previous concepts focussed around biochemical pathways reporting directionally discordant pleiotropy to discover metabolic bottlenecks ^74^.

Disturbance in metabolism or rearrangements thereof are a hallmark of many diseases, including those not classically considered as ‘metabolic’, such as eye disorders^2^, but whether these are pathways for prevention or intervention rather than a consequence of the disease remains often elusive in humans. We demonstrated considerable overlap between mQTLs with disease risk loci, including rare-to-common allelic series that can reveal unknown effector genes. However, many such ‘locus effects’ were characterised by nonspecific pleiotropy, implicating the plasma metabolite as a bystander rather than cause of the disease. This observation aligns with the relatively few notable exceptions, such as HDL particle characteristics and CAD, from two-sample MR analyses that contrasted the broad spectrum of observed disease-associations described for the same NMR platform^75^. These observations might be best explained by the concept of metabolic flexibility, which includes built-in redundance in key pathways to combat various intrinsic and extrinsic perturbations.

An important distinction of our study compared to most previous efforts was the availability of highly standardized measurements in a well-designed single large cohort, mitigating influences of preanalytical variables and enabling analyses of even ultra-rare variants. However, this also meant that we had little opportunity to investigate the influence of different states of metabolism on our genetic results (such as an overnight fast) or investigate robustness of findings in different enviroments or at scale in other ancestries. For example, UKB participants were not asked to fast overnight prior to their baseline visit, which has been shown to impact genetic findings^3^. Consequently, sentinel variants capture relatively little variance (0.57% - 1.07%) in circulating ketone body concentrations that are highly dependent on the time since last food consumption. Other limitations included the sensitivity and coverage of the ^1^H-NMR platform, and future efforts are likely to reveal more diverse phenotypic consequences of genetically constrained flexibility of human metabolism. Another technical aspect to consider in the interpretation of our results is the indirect nature of ^1^H-NMR derived measurements of certain analytes, including apolipoproteins, that may have no longer be reliable in the presence of rare damaging variants that change the properties of apolipoproteins as observed for ApoA1.

## Data Availability

All individual-level data is publicly available to bona fide researchers from the UK Biobank (https://www.ukbiobank.ac.uk/). Summary statistics are available through the GWAS catalog (see Github: https://github.com/comp-med/ukb-mgwas)

https://www.ebi.ac.uk/gwas/

## Acknowledgements

The authors acknowledge the Scientific Computing of the IT Division at the Charité - Universitätsmedizin Berlin for providing computational resources that have contributed to the research results reported in this paper (https://www.charite.de/en/research/research_support_services/research_infrastructure/science_it/#c30646061). We acknowledge Nightingale Health Plc for access to the UK Biobank NMR biomarker data. We are deeply grateful to the participants, investigators and teams of the UKB and FinnGen studies. We thank Benjamin Wild for assistance in data processing and helpful discussions. This work was supported by DZHK (German Centre for Cardiovascular Research) and BMBF (German Ministry of Education and Research) grants to C.L. and co-funded by a European Union grant to M.P (ERC, GenDrug, 101116072). Views and opinions expressed are however those of the author(s) only and do not necessarily reflect those of the European Union or the European Research Council. Neither the European Union nor the granting authority can be held responsible for them. The funders had no role in study design, data collection and analysis, decision to publish or preparation of the manuscript.

## Author Contributions Statement

Conceptualization: MZ, MP, CL

Data curation/Software: MZ, CB, YS, LK, MP

Formal Analysis: MZ, CB, YS, LK, AW, MP

Methodology: MZ, CB, YS, LK, MK, AW, MP, CL

Visualization: MZ, CB, LK, AW, MP, CL

Funding acquisition: CL, MP

Project administration: CL

Supervision: MP, CL

Writing – original draft: MZ, CB, YS, MP, CL

Writing – review & editing: MZ, CB, YS, LK, MK, FK, MM, AW, MP, CL

## Data availability

All individual-level data is publicly available to bona fide researchers from the UK Biobank (https://www.ukbiobank.ac.uk/). Full summary statistics for all analyses are publicly available through the NHGRI-EBI GWAS Catalogue (see Github repository for GWAS Catalogue identifiers).

## Code availability

Code for the main analyses is freely available on Github (https://github.com/comp-med/ukb-mgwas) and permanently archived on Zenodo (doi.org/10.5281/zenodo.14716599).

## Methods

### Study design

UK Biobank is a prospective cohort study from the UK that contains more than 500,000 volunteers between 40 and 69 years of age at inclusion. The study design, sample characteristics, and genotype data have been described elsewhere^76,77^. The UKBB was approved by the National Research Ethics Service Committee North West Multi-Centre Haydock and all study procedures were performed in accordance with the World Medical Association Declaration of Helsinki ethical principles for medical research. We included 460,036 individuals across the three major ancestries in the UK Biobank in our analyses for whom inclusion criteria (given consent to further usage of the data, availability of genetic data, and passed quality control of genetic data) applied. Data from the UKBB were linked to death registries and hospital episode statistics (HES). We used the ancestry assignments as defined by the pan-UKB^78^, and further made an effort to assign unclassified individuals to their respective ancestries based on a k-nearest neighbour approach using genetic principal components. All analyses were conducted under UKBB application 44448 and 30418.

### Metabolomic measurements

Up to 249 targeted metabolomic measurements were quantified using the Nightingale NMR platform in human EDTA plasma samples. Detailed experimental procedures for the NMR platform are described elsewhere^75,79^. The NMR platform covers a wide range of metabolic biomarkers, including lipoprotein lipids, fatty acids as well as small molecules such as amino acids, ketone bodies and glycolysis metabolites. All metabolites are quantified in molar concentration units. We combine here three data releases that cover the full breadth of the UKBB. Metabolomics data was available for 482276 individuals, including 19699 samples with data from both the baseline and repeat visit.

Metabolites were reliably detected, with only one biomarker over 2.5% missingness in releases 1/2 (creatinine) and release 3 (3-Hydroxybutyrate). 98% of the samples had < 5% missingness over all biomarkers in releases 1/2 and release 3. We used the *ukbnmr*^80^ R package (v2.2, R v4.3.2) for quality control and removal of technical variation in the NMR data. This includes technical confounders such as sample preparation time, shipping plate well, spectrometer effects, time drift within spectrometers and outlier plates.

We removed samples that were flagged by Nightingale for poor quality and used the MICE (Multivariate Imputation by Chained Equations)^81^ R package to impute the remaining dataset. In total, we imputed 0.16% and 0.17% of data in releases 1/2 and release 3, respectively.

We observed overall good consistency with the overlapping routine blood biomarkers previously measured in the same cohort (median R^2^: 0.9, range: 0.62 – 0.94) (**Supplementary Fig. 14**).

### Adjustment of metabolomic data for medication use

We sought to adjust the NMR data for medication use, especially cholesterol-lowering medication to avoid false positive results driven by medication use in downstream genetic analyses. For male and female participants separately, we fitted linear models to quantify the impact of 6 drug categories on each NMR phenotype: cholesterol-lowering medicine, blood pressure medication, diabetic medication including Metformin usage, oral contraceptive pill or minipill (female only), hormone replacement therapy (female only) (UKB fields 6177 and 6153) (**Supplementary Fig. 15, Supplementary Table 18**).

We used data from individuals with both baseline and repeat assessment metabolic data available and estimated the effect of medication in individuals that did not take any drugs at the time of the baseline visit (N = 6,312 male, N = 6,713 female participants). We fitted a linear model predicting the follow-up metabolic data from baseline metabolic data, sex, age and medication use:

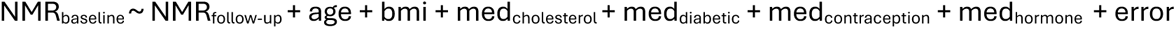

We note that the sample sizes for diabetic medication (N_male_ = 45, N_female_ = 29), oral contraceptive medication (N = 27) and hormone replacement therapy (N=148) were too small to reliably estimate any effects. Effect estimates for diabetic medication were correlated to estimates for cholesterol-lowering medicine. The effect estimates for blood pressure medication were minimal across the phenotypes. We considered thus only the impact of cholesterol-lowering medicine and corrected the metabolic data in a sex-specific manner.

### Genotyping and GWAS analyses

GWAS was performed on 249 metabolic traits measured by the NMR platform on European (n = 434,646), British-Asian (n = 8,796) and British-African participants (n=6,573) that had complete phenotypic, covariate and genetic information available. We performed GWAS under the additive model using REGENIE (v3.2.5)^82^ that employs a two-step procedure to account for population structure. We derived a set of high-quality genotyped variants per population by applying following filters: (MAF > 1%, MAC > 100, missingness rate < 10%, pHWE > 10^-15). Further, linkage disequilibrium pruning was performed using a 1000 kb window, shifting by 100 variants and removing variants with LD(r2) > 0.8. We used these variants as input for the first step of REGENIE, to generate individual trait predictions using the leave-one-chromosome-out scheme. These predictions are used in the second step where individual variants are tested. Models were adjusted for age, sex and the first ten genetic principal components. We tested variants with a minor allele frequency > 0.5%, amounting to 11.5M variants in European individuals, 11.5M variants in British-Asian individuals and 19.3M variants in British-African individuals.

For initial discovery, we performed a meta-analysis across the three ancestral groups using METAL^83^. We required variants to be present in at least two ancestral groups. To declare significance, we considered a stringent p-value threshold (2.0 x 10^-10^) by dividing the standard genome-wide threshold by the number of metabolic phenotypes (5.0 x 10^-8^/ 249).

We tested our results for genomic inflation and calculated the SNP-based heritability using LD-score regression (LDSC)^84^ (**Supplementary Table 19**).

### Regional clumping and fine-mapping

We used regional clumping (±500kb) around sentinel variants from the analyses including White European samples to select independent genomic regions associated with a metabolic phenotype and collapsed neighbouring regions using BEDtools (v2.30.0). We treated the extended MHC region (chr6:25.5-34.0Mb) as one region.

Within each region of interest, excluding the MHC region, we performed statistical fine-mapping for all phenotypes associated with that region using the ‘Sum of single effects’ model (SuSiE) implemented in the *susieR* (v0.12.35) R package^85^. Briefly, SuSiE employs a Bayesian framework for variable selection in a multiple regression problem with the aim to identify sets of independent variants each of which likely contains the true causally underlying genetic variant. We implemented the workflow using default prior and parameter settings, apart from the minimum absolute correlation, which we set to 0.1. Since SuSiE is implemented in a linear regression framework, we used the GWAS summary statistics with a matching correlation matrix of dosage genotypes instead of individual level data to implement fine-mapping (*susie_rss()*) as recommended by the authors^85^.

Within a given region, a phenotype can be associated to multiple, independent genetic loci. To determine the appropriate number of credible sets, we iterated over the maximum credible sets parameter in *susieR* from two to ten, thus generating fine-mapped results constrained to a range of maximum number of credible sets. For each collection of credible sets, we pruned sets where the lead variant was correlated to the lead variant of other credible sets (R^2^ > 0.25). After pruning, we considered the fine-mapped results constrained to the largest number of credible sets that still contained one or more credible sets.

We performed several sensitivity analyses by computing joint models per locus – phenotype combination. We obtained all lead variants across the credible sets provided by SuSiE based on posterior inclusion probability and fitted a single linear model, jointly modelling the effect of all distinct credible sets in the locus for a given phenotype. Subsequently, we retained only credible sets where the lead variant reached genome-wide significance (p = 5.0 x 10^-8^) in both marginal and joint statistics. Furthermore, we ensured the estimated coefficients were directionally concordant and of similar magnitude between joint and marginal models (± 25%). Linear models were implemented in R using the *glm()* function and used only unrelated white-European participants and the same set of covariates as described above.

Finally, we used LD-clumping (r^2^ > 0.6) to identify credible sets shared across metabolic phenotypes.

We computed the correlation matrix with LDscore v2.0 using genetic data from 50,000 randomly selected, unrelated White European UKB participants. In situations where SuSiE did not deliver a credible set, we used the Wakefield approximation^86^ to compute 95%-credible sets.

### Multi-ancestry finemapping

We aimed to use ancestrally diverse genetic data to refine the credible sets identified in the White European analyses. For 35577 credible sets that contained two or more variants in the European analyses, we checked for evidence of a genetic signal in the British-African and British-Central/South Asian ancestries (P < 1.0 x 10^-4^) at the same locus (±25kb on either side of the credible set). After these filters, we considered 6979 credible sets for finemapping across ancestries using MultiSuSiE^87^. We considered only quality-controlled variants that were prevalent in all populations (MAF>0.5%) and used the posterior inclusions probabilities from the European analyses as priors. LD matrices were calculated from a random subset of 50,000 White European participants for Europeans, and using all available individuals for the British-African and British-Central/South Asian ancestries.

### Replication of genetic associations

We replicated our trans-ancestral genetic signals using two independent studies: i) the so-far largest published mGWAS^3^, and ii) a parallel effort using overlapping UK Biobank data^9^, both using the same NMR platform. We considered a set of metabolic traits that were directly measured by the NMR platform and not inferred from other traits to avoid multiplicative errors in these more sensitive phenotypes. In total, we were able to match 144 (Karjalainen et al) and 169 (Tambets et al) metabolic traits, for which we compared sentinel variants that passed metabolome-adjusted, genome-wide significance in our trans-ancestral meta-analysis and that overlapped between the studies.

### Sex-specific genetic analyses

To assess whether our genetic analyses were driven by sex differences and whether our results were transferrable to both sexes, we performed sex-stratified GWAS within the largest ancestry (EUR). We defined ‘female’ and ‘male’ sex including participants where the recorded sex and sex chromosomes aligned (XX for females and XY for males). The recorded sex was self-reported, and it was not possible to distinguish sex from gender. We acknowledge the importance of distinguishing between sex and gender in research and that chromosomal make-up does not always align with self-identified gender. In total, phenotypic, covariate and genetic data was fully available for 198,796 males and 235,850 females. GWASs were performed using REGENIE as described above. Per metabolic trait, we meta-analysed the sex-stratified results using the inverse-variance weighted model in METAL (v2020-05-05)^83^ and finally clumped results on the heterogeneity p-value using plink (v2.00)^88^ (--clump-p1 5 x 10^-8^, --clump-r2 0, --clump-kb 2500). We considered loci putatively sex-differential if the meta-analysis heterogeneity p-values were genome-wide significant (p < 5 x 10^-8^). We performed additional sensitivity analyses on the putatively sex-differential loci by assessing the influence of covariates confounded by sex (BMI, tobacco usage, alcohol consumption, lipid-lowering medication and diabetic medication). To properly model all gene by environment interactions^89^, we fitted the following model per clump lead variant and associated metabolic phenotype, including the same set of covariates used in the original GWAS:

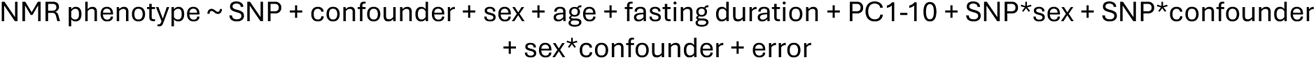

### Causal gene assignment

To assign candidate genes for all metabolite-QTLs residing outside the MHC region, we first collected annotations for each genetic variant or proxies thereof (r^2^ > 0.6), including 1) distance to the gene body and 2) putative functional consequences based on the Variant Effect Predictor (VEP) tool offered by Ensembl. We further collated up to 10 closest genes within a 2 Mb window and subsequent gene features such as: 1) eQTL evidence for a given variant-gene pair for each tissue available in the eQTL Catalogue release 7^90^, 2) evidence of being annotated as metabolic in the MGI or Orphanet databases as defined in ProGem^25^, 3) evidence of being listed in the OMIM database^43^ 4) and evidence of being an already assigned drug target in Open Targets^91^ clinical stage III and IV.

With no universally accepted standard for variant-to-gene assignments, we relied on prior biological and genomic information to create three sets of “putative true positive” (PTP) set: 1) genes annotated as part of a cholesterol pathway in the KEGG^92^ or REACTOME^93^ database (n=6791, 722 unique SNPs), 2) genes annotated as part of a lipid pathway (n=5670, 603 unique SNPs) and 3) genes annotated as part of an amino acid-related pathway (n=8349, 895 unique SNPs). We used all fine-mapped SNPs associated with metabolites classified in the respective NMR metabolite class (Cholesterol: Cholesterol, Cholesteryl esters, Free cholesterol; Lipid: Total lipids, other lipids, Relative lipid concentration, Phospholipids); Amino Acid: Amino acid) in the PTP set and used overlapping SNPs in only one PTP set. The dataset was split in a 7:3 ratio to obtain training and test sets without overlapping variants. We trained a Random Forest classifier using 5-fold cross-validation with implemented subsampling to account for the unbalanced datasets. The implementation was carried out using python scikit-learn v1.4.1. We used the balanced accuracy score to choose the best-performing forest from each training set. Subsequently, we used the best-performing Random Forest classifiers from each PTP set to assign candidate scores for all putative effector genes across the entire set of metabolite-QTLs. We then calculated the median score of these classifiers and selected the highest-scoring gene as the assigned gene for the variant. Within each PTP set, we omitted features used to define true positive sets. Each of the three classifiers exhibited consistent performance with a mean ROC AUC of 0.80 and a mean balanced accuracy score of 0.69 (**Supplementary Fig. 16**).

To provide another layer of evidence for assignment of causal genes at metabolic loci, we performed cis-colocalisation with protein targets measured in the independent Fenland study^28^. Cis (e.g. gene body ± 500kb) summary statistics were preprocessed using MungeSumStats^94^. To relax the single causal variant assumption, we employed a colocalization approach where we fine-mapped all traits with SuSiE and then performed colocalization among all credible sets using functionality of the ‘coloc’ (v5.2.3)^95,96^ and ‘susieR’ (v0.12.35)^85^ R packages. For this, we set the prior probability that a SNP is associated with both traits to 5×10^-6^ and restricted the maximum number of credible sets for the outcome data to 5^95^.

### Tissue enrichment of metabolic loci

We tested whether genes proximal to metabolic loci and assigned effector genes were enriched in tissue compartments by leveraging data from the Human Protein Atlas^97^. Specifically, we used a two-sided Fisher’s test whether metabolic genes were enriched among tissue-specific genes (tissue-enriched or tissue-enhanced as defined by the Protein Atlas) against all protein-coding genes as background.

### Pleiotropy assignment and overlap with the GWAS catalog

To assign modes of pleiotropy for each mQTL, we first clumped lead credible set variants across NMR measures by LD, collating variants with r^2^≥0.6 as a single signal, referred to hereafter as ‘mQTL. This was done based on dosage files of all unrelated White European UKB participants and implemented with the *igraph* (v.2.0.1.1) package in R. For each mQTL, we then computed all possible Pearson correlation coefficients among associated NMR measures. To classify each mQTL-group, generated two metrics: 1) the 25^th^ percentile of all correlations among associated NMR measures, and 2) the Pearson correlation coefficient between the association strengths for each measure (-log_10_(p-value) and its correlation coefficient with the most strongly associated measure within the mQTL. The latter is a measure to what extend the association between NMR measures at a given locus (‘pleiotropy’) can be explained by being correlated with the most proximal associated measure. Based on opposing those two measures for all mQTLs we opted to threshold each at 0.6 to define the following five groups: 1) ‘specific’ mQTLs associated with only ≤3 highly correlated NMR measures (rho≥0.6), 2) ‘pathway pleiotropic’ mQTLs associated with highly correlated NMR measures (rho≥0.6) that also followed the described association pattern (rho≥0.6), 3) ‘proportional pleiotropic’ mQTL groups associated with, in part, uncorrelated NMR measures but highly correlated association statistics (rho≥0.6), 4) ‘disproportional pleiotropic’ mQTLs associated with highly correlated NMR measures (rho≥0.6), but without evidence that this translated into a correlation of association statistics (rho<0.6), and 5) all remaining mQTLs as ‘unspecific pleiotropic’ groups.

To quantify the extent to which our pleiotropy assignment extends beyond the NMR measured analysed here, we intersected mQTLs and proxies thereof with results reported in the GWAS catalog (download: 20/05/2024). We first pruned GWAS catalog entries for those with mapped traits (to minimize double counting), results that met genome-wide significance (p<5×10^-8^) and had location information available. We further dropped results similar to NMR measures based on broad EFO terms (e.g., EFO:0005105 and child terms indicating ‘lipid or lipoprotein measurement’). To further account for traits mapping to similar categories, we iteratively traced back mapped EFO terms to broader parent terms. We finally classified mQTLs to be ‘specific’ in the GWAS catalog, if they associated with less than five parent EFO-terms and ‘unspecific’ otherwise. This information was primarily used to define instruments for Mendelian randomization analysis.

### Integration of metabolomic measurements with cardiovascular endpoints

We next aimed to utilize the mQTLs to investigate the shared genetic basis of the 249 NMR and 25 selected cardiovascular disease (CVD) traits. We utilized public databases (GWAS Catalog, openGWAS, CVD-KP) to collect CVD data comprising the largest currently publicly available GWASs on coronary artery disease and myocardial infarction, angina pectoris, aortic aneurysm, heart failure and stroke, peripheral arterial disease including 2-5 subtypes for each phenotype. An additional 10 CVD traits had no subtype data available (**Supplementary Table 13**) Data was harmonized and if necessary, lifted over to GRCh37 using the ‘MungeSumstats’ (v1.13.2) R package^94^. We queried mQTL lead variants and proxies in strong LD (r^2^>0.8; LD backbone based on UK Biobank, as described above) of each NMR trait in each region and corresponding summary statistics for each CVD trait.

To investigate ‘variant’ effects on NMR metabolite concentrations and CVD outcomes, we performed statistical colocalization screens for all combinations of the NMR traits in regions with at least one credible set and CVD traits with matching summary statistics^98^. We applied statistical colocalization as described before (see ‘Causal gene assignment’).

To estimate ‘level’ effects of NMR metabolite concentrations on CVD outcomes, we performed Mendelian Randomization analysis using the ‘TwoSampleMR’ package (v0.5.1), implementing the inverse-variance weighted and the MR-Egger methods. We used all 249 NMR metabolites as exposure variables, the 25 CVDs as outcome variables and assessed separately four sets of instruments: 1) sentinel variants, 2) lead credible set variants, 3) lead credible set variants restricted for molecular pleiotropy (e.g. ‘pathway pleiotropy’) and 4) lead credible set variants restricted for both molecular and phenotypic pleiotropy. We used the Wald ratio method to estimate the effect of NMR concentrations on CVD outcomes using only single genetic variants^99^. We used MR Egger to test for evidence of a pleiotropic association, an intercept p-value of p> 0.0001 indicating evidence of no pleiotropy and checked for concordance between the effect estimates of IVW-MR, MR-Egger and single genetic variant MR. We controlled the false discovery rate (FDR) at FDR=5% ^100^. To further limit the possible extend of pleiotropic associations, we only reported ‘level effects’ passing these filters in the variant sets 2-4, prioritizing the association in the more stringent variant set.

The overlap of ‘locus effects’ showing no ‘disproportional pleiotropy’ according to the section **Pleiotropy assignment and overlap with the GWAS catalog** as well as a significant single variant MR (FDR=5%) and ‘level effects’ calculated from metabolite-specific or metabolite- and phenome-specific variants was used to identify gene-metabolite pairs associated with cardiovascular disease risk independent of LDL-metabolism. We considered loci as independent from LDL-metabolism if they did not associate with clinical LDL-cholesterol at the locus with p < 2.0 x 10^-10^ and the effect estimate of any variant on clinical LDL-C ranked upwards the 80^th^ percentile of all effect estimates at the locus.

### Rare Variant Analyses with whole exome sequencing data

#### Whole Exome Sequencing data QC

An in-depth description of whole exome sequencing, including experimental details, variant calling, and standard quality control measures for the UK Biobank, has been extensively reported by Backman et al.^101^. We performed additional quality control (QC) steps at the UKB Research Analysis Platform (RAP; https://ukbiobank.dnanexus.com/).

We employed bcftools (v1.15.1) to process population-level Variant Call Format (pVCF) files. Initially, we normalised the data using the reference sequence GRCh38 build, followed by splitting multi-allelic variants. Subsequently, we conducted QC on these variants using a set of parameters outlined below to filter high-quality variants for downstream genetic analyses. Genotypes for SNPs were set to missing if the read depth was less than 7 (or less than 10 for INDELs) or if the genotype quality was below 20. Furthermore, we excluded variants if the allele balance (AB) was less than 0.25 or greater than 0.8 in heterozygous carriers.

Finally, we computed the missingness rate for each variant and excluded those with missing values in over 50% of the participants.

#### Variant Annotation and Gene burden Masks

Variants were annotated using ENSEMBL Variant Effect Predictor (VEP)^102^ (v106.1) with the most severe consequence for each variant chosen across all protein-coding transcripts. We further utilized additional plugins REVEL^103^, CADD v1.6^104^, and LOFTEE^105^, for variant annotation. Based on these scores we defined six partially overlapping variant masks: 1) high-confidence predicted loss-of-function (pLOF, based on LOFTEE and includes stop-gained, splice site disrupting, and frameshift variants), 2) any pLOF assigned high impact by VEP, 3) pLOF and high-impact missense variants (CADD score > 20 oe REVEL score > 0.5), 4) pLOF and any missense variants, 5) only high-impact variants, and 6) any missense variants but not pLOF. We tested synonymous variants separately as a negative control. We tested each mask in different minor allele frequency bins, using 0.5% and 0.005% as thresholds.

We performed rare variant association testing (RVAT) using WES data across 249 quantitative NMR phenotypes using REGENIE (v3.1.1) via the DNAnexus Swiss Army Knife tool (v4.9.1). Similar to common variant GWASs, we used a two-step approach by REGENIE. However, we additionally generated step1 LOCO files with and without adjusting for common signals via a polygenic score (PGS) in the RVAT models per phenotype. In practice, we computed a PGS for each phenotype using effect sizes of lead variants from the GWAS summary statistics and corresponding dosages of variants from imputed data. All RVAT models were then adjusted for PGS in addition to age, biological sex, fasting duration and the first ten genetic PCs. We first performed aggregated gene burden testing across for 19,026 genes using a set of masks as defined above. For the gene burden testing we used aggregated Cauchy association test (ACAT) to estimate a p-value for each gene across all combinations of masks and allele frequency bins. ACAT first computes p-values for all sets defined by various masks within a gene and then takes these p-values as input to compute one p-value for the respective gene via a well approximated Cauchy distribution.

We have also performed single variant association testing for exonic variants commonly referred to as exome wide association study (ExWAS). For the ExWAS, we only tested variants with MAC >5 and reported results for variants with a MAF < 0.0005. We have performed these analyses in individuals of White Europeans, British African and British South Asian ancestry.

We considered findings as robust, if they passed multiple testing corrected statistical significance (gene burden: p<1.2×10^-8^ [corrected for the number of genes x number of traits]; ExWAS: p<2.0 x 10^-10^ [same as for common variant GWAS, conventional genome-wide significance corrected for the number of traits]) in both the model with and without adjusting for the common variant PGS and effect sizes did not differ by more than 20% between these models, since this might otherwise indicate that rare variant findings cannot clearly distinguished from common variant effects.

#### Phenotype definition

To systematically test for phenotypic consequences of genes identified through rare variant analysis, we collated 626 disease entities following previous work^1^ by aggregating information from self-report, hospital episode statistics, death certificates, and primary care data (45% of the UKB population). Each of the disease entities had at least one common variant finding passing statistical significance, and we employed a similar analysis workflow using REGENIE as described for NMR measures but using logistic regression with saddle point approximation.

#### Integration of OMIM (Online Mendelian Inheritance in Man)

We downloaded the OMIM gene – disease list (09/11/2023) and kept 7,327 unique entries after filtering for gene entries with high confidence (level 3). We computed the enrichment of genes associated with any NMR measure from rare variant or gene burden analysis against a background of 19,989 protein coding genes using Fisher’s exact test.

### Mendelian Randomisation analyses

We performed a phenome-wide Mendelian Randomisation screen using outcome summary statistics from the independent FinnGenn^68^ cohort, release 11 (June 2024). We assessed only outcomes with genome-wide significant signals (5 x 10^-8^), yielding 1394 phenotypic outcomes. We selected 21 non-lipid NMR biomarkers as exposure variables and assessed separately four sets of instruments as described previously for the cardiovascular Mendelian Randomisation analyses. We included two well-characterized lipid biomarkers (LDL-C and ApoB) as positive controls in the MR analyses.

We performed MR using the TwoSampleMR package (v0.5.1), implementing the inverse-variance weighted and the MR-Egger methods. We discarded results with MR Egger p < 0.001, Cochran’s Q p-value < 1.0 x 10^-6^ and results where the estimated effect was directionally discordant between the IVW and Egger methods.

## References

1. Surendran, P. et al. Rare and common genetic determinants of metabolic individuality and their effects on human health. Nat. Med. 28, 2321–2332 (2022).

2. Lotta, L. A. et al. A cross-platform approach identifies genetic regulators of human metabolism and health. Nat. Genet. 53, 54–64 (2021).

3. Karjalainen, M. K. et al. Genome-wide characterization of circulating metabolic biomarkers. Nature 628, 130–138 (2024).

4. Kettunen, J. et al. Genome-wide association study identifies multiple loci influencing human serum metabolite levels. Nat. Genet. 44, 269–276 (2012).

5. Chen, Y. et al. Genomic atlas of the plasma metabolome prioritizes metabolites implicated in human diseases. Nat. Genet. 55, 44–53 (2023).

6. Nag, A. et al. Effects of protein-coding variants on blood metabolite measurements and clinical biomarkers in the UK Biobank. Am. J. Hum. Genet. 110, 487–498 (2023).

7. Long, T. et al. Whole-genome sequencing identifies common-to-rare variants associated with human blood metabolites. Nat. Genet. 49, 568–578 (2017).

8. Shin, S.-Y. et al. An atlas of genetic influences on human blood metabolites. Nat. Genet. 46, 543–550 (2014).

9. Tambets, R. et al. Genome-wide association study for circulating metabolites in 619,372 individuals. medRxiv (2024) doi:10.1101/2024.10.15.24315557.

10. Yin, X. et al. Genome-wide association studies of metabolites in Finnish men identify disease-relevant loci. Nat. Commun. 13, 1644 (2022).

11. Van Der Meer, D. et al. Pleiotropic and sex-specific genetic architecture of circulating metabolic markers. medRxiv (2024) doi:10.1101/2024.07.30.24311254.

12. Khan, A. et al. Metabolic gene function discovery platform GeneMAP identifies SLC25A48 as necessary for mitochondrial choline import. Nat. Genet. (2024) doi:10.1038/s41588-024-01827-2.

13. Schlosser, P. et al. Genetic studies of paired metabolomes reveal enzymatic and transport processes at the interface of plasma and urine. Nat. Genet. 55, 995–1008 (2023).

14. Love-Gregory, L. et al. Variants in the CD36 gene associate with the metabolic syndrome and high-density lipoprotein cholesterol. Hum. Mol. Genet. 17, 1695–1704 (2008).

15. Adiyaman, S. C. et al. Congenital generalized lipodystrophy type 4 due to a novel PTRF/CAVIN1 pathogenic variant in a child: effects of metreleptin substitution. J. Pediatr. Endocrinol. Metab. JPEM 35, 946–952 (2022).

16. Mauvais-Jarvis, F. Sex differences in energy metabolism: natural selection, mechanisms and consequences. Nat. Rev. Nephrol. 20, 56–69 (2024).

17. Gerdts, E. C Regitz-Zagrosek, V. Sex differences in cardiometabolic disorders. Nat. Med. 25, 1657–1666 (2019).

18. Wittemans, L. B. L. et al. Assessing the causal association of glycine with risk of cardio-metabolic diseases. Nat. Commun. 10, 1060 (2019).

19. Mittelstrass, K. et al. Discovery of sexual dimorphisms in metabolic and genetic biomarkers. PLoS Genet. 7, e1002215 (2011).

20. Koprulu, M. et al. Similar and different: systematic investigation of proteogenomic variation between sexes and its relevance for human diseases. medRxiv (2024) doi:10.1101/2024.02.16.24302936.

21. Michos, E. D., McEvoy, J. W. & Blumenthal, R. S. Lipid Management for the Prevention of Atherosclerotic Cardiovascular Disease. N. Engl. J. Med. 381, 1557– 1567 (2019).

22. Graham, S. E. et al. The power of genetic diversity in genome-wide association studies of lipids. Nature 600, 675–679 (2021).

23. BasuRay, S., Wang, Y., Smagris, E., Cohen, J. C. & Hobbs, H. H. Accumulation of PNPLA3 on lipid droplets is the basis of associated hepatic steatosis. Proc. Natl. Acad. Sci. U. S. A. 116, 9521–9526 (2019).

24. Johnson, S. M. et al. PNPLA3 is a triglyceride lipase that mobilizes polyunsaturated fatty acids to facilitate hepatic secretion of large-sized very low-density lipoprotein. Nat. Commun. 15, 4847 (2024).

25. Stacey, D. et al. ProGeM: a framework for the prioritization of candidate causal genes at molecular quantitative trait loci. Nucleic Acids Res. 47, e3 (2019).

26. Vujkovic, M. et al. Discovery of 318 new risk loci for type 2 diabetes and related vascular outcomes among 1.4 million participants in a multi-ancestry meta-analysis. Nat. Genet. 52, 680–691 (2020).

27. Donaldson, J. G. & Jackson, C. L. ARF family G proteins and their regulators: roles in membrane transport, development and disease. Nat. Rev. Mol. Cell Biol. 12, 362– 375 (2011).

28. Pietzner, M. et al. Mapping the proteo-genomic convergence of human diseases. Science 374, eabj1541 (2021).

29. Pellegrinelli, V. et al. Dysregulation of macrophage PEPD in obesity determines adipose tissue fibro-inflammation and insulin resistance. Nat. Metab. 4, 476–494 (2022).

30. Wang, Q., et al. Metabolic profiling of angiopoietin-like protein 3 and 4 inhibition: a drug-target Mendelian randomization analysis. Eur. Heart J. 42, 1160–1169 (2021).

31. Hindy, G. et al. Rare coding variants in 35 genes associate with circulating lipid levels-A multi-ancestry analysis of 170,000 exomes. Am. J. Hum. Genet. 109, 81–96 (2022).

32. Sjouke, B., Balak, D. M. W., Beuers, U., Ratziu, V. C Stroes, E. S. G. Is mipomersen ready for clinical implementation? A transatlantic dilemma. Curr. Opin. Lipidol. 24, 301–306 (2013).

33. Spracklen, C. N. et al. Identification of type 2 diabetes loci in 433,540 East Asian individuals. Nature 582, 240–245 (2020).

34. Rhee, E. P. et al. An exome array study of the plasma metabolome. Nat. Commun. 7, 12360 (2016).

35. Chen, X., Gu, X. & Zhang, H. Sidt2 regulates hepatocellular lipid metabolism through autophagy. J. Lipid Res. 59, 404–415 (2018).

36. Sampieri, A., Asanov, A., Méndez-Acevedo, K. M. & Vaca, L. SIDT2 Associates with Apolipoprotein A1 (ApoA1) and Facilitates ApoA1 Secretion in Hepatocytes. Cells 12, 2353 (2023).

37. Jaiswal, S. et al. Clonal Hematopoiesis and Risk of Atherosclerotic Cardiovascular Disease. N. Engl. J. Med. 377, 111–121 (2017).

38. Weiner, D. J. et al. Polygenic architecture of rare coding variation across 394,783 exomes. Nature 614, 492–499 (2023).

39. Szeri, F. et al. The membrane protein ANKH is crucial for bone mechanical performance by mediating cellular export of citrate and ATP. PLoS Genet. 16, e1008884 (2020).

40. Chroni, A. & Kardassis, D. HDL Dysfunction Caused by Mutations in apoA-I and Other Genes that are Critical for HDL Biogenesis and Remodeling. Curr. Med. Chem. 26, 1544–1575 (2019).

41. Tilly-Kiesi, M. et al. ApoA-I_Helsinki_ (Lys_107_ →0) Associated With Reduced HDL Cholesterol and LpA-I:A-II Deficiency. Arterioscler. Thromb. Vasc. Biol. 15, 1294–1306 (1995).

42. Zanoni, P. & Von Eckardstein, A. Inborn errors of apolipoprotein A-I metabolism: implications for disease, research and development. Curr. Opin. Lipidol. 31, 62–70 (2020).

43. Amberger, J. S., Bocchini, C. A., Schiettecatte, F., Scott, A. F. & Hamosh, A. OMIM.org: Online Mendelian Inheritance in Man (OMIM®), an online catalog of human genes and genetic disorders. Nucleic Acids Res. 43, D789–D798 (2015).

44. Horiki, M. et al. Smad6/Smurf1 overexpression in cartilage delays chondrocyte hypertrophy and causes dwarfism with osteopenia. J. Cell Biol. 165, 433–445 (2004).

45. Zhang, F., Sodroski, C., Cha, H., Li, Q. C Liang, T. J. Infection of Hepatocytes With HCV Increases Cell Surface Levels of Heparan Sulfate Proteoglycans, Uptake of Cholesterol and Lipoprotein, and Virus Entry by Up-regulating SMAD6 and SMAD7. Gastroenterology 152, 257–270.e7 (2017).

46. Plenge, R. M., Scolnick, E. M. & Altshuler, D. Validating therapeutic targets through human genetics. Nat. Rev. Drug Discov. 12, 581–594 (2013).

47. Hoogeveen, R. C. & Ballantyne, C. M. Residual Cardiovascular Risk at Low LDL: Remnants, Lipoprotein(a), and Inflammation. Clin. Chem. 67, 143–153 (2021).

48. Aragam, K. G. et al. Discovery and systematic characterization of risk variants and genes for coronary artery disease in over a million participants. Nat. Genet. 54, 1803–1815 (2022).

49. Sakaue, S. et al. A cross-population atlas of genetic associations for 220 human phenotypes. Nat. Genet. 53, 1415–1424 (2021).

50. Roychowdhury, T. et al. Genome-wide association meta-analysis identifies risk loci for abdominal aortic aneurysm and highlights PCSK9 as a therapeutic target. Nat. Genet. 55, 1831–1842 (2023).

51. Roychowdhury, T. et al. Regulatory variants in TCF7L2 are associated with thoracic aortic aneurysm. Am. J. Hum. Genet. 108, 1578–1589 (2021).

52. Miyazawa, K. et al. Cross-ancestry genome-wide analysis of atrial fibrillation unveils disease biology and enables cardioembolic risk prediction. Nat. Genet. 55, 187–197 (2023).

53. Yu Chen, H., et al. Dyslipidemia, inflammation, calcification, and adiposity in aortic stenosis: a genome-wide study. Eur. Heart J. 44, 1927–1939 (2023).

54. Zhou, W. et al. Global Biobank Meta-analysis Initiative: Powering genetic discovery across human disease. Cell Genomics 2, 100192 (2022).

55. Kavousi, M. et al. Multi-ancestry genome-wide study identifies effector genes and druggable pathways for coronary artery calcification. Nat. Genet. 55, 1651–1664 (2023).

56. Henry, A. et al. Mapping the aetiological foundations of the heart failure spectrum using human genetics. medRxiv (2023) doi:10.1101/2023.10.01.23296379.

57. Ishigaki, K. et al. Large-scale genome-wide association study in a Japanese population identifies novel susceptibility loci across different diseases. Nat. Genet. 52, 669–679 (2020).

58. Mishra, A. et al. Stroke genetics informs drug discovery and risk prediction across ancestries. Nature 611, 115–123 (2022).

59. Roselli, C. et al. Genome-wide association study reveals novel genetic loci: a new polygenic risk score for mitral valve prolapse. Eur. Heart J. 43, 1668–1680 (2022).

60. Hartiala, J. A. et al. Genome-wide analysis identifies novel susceptibility loci for myocardial infarction. Eur. Heart J. 42, 919–933 (2021).

61. van Zuydam, N. R. et al. Genome-Wide Association Study of Peripheral Artery Disease. Circ. Genomic Precis. Med. 14, e002862 (2021).

62. Adlam, D. et al. Genome-wide association meta-analysis of spontaneous coronary artery dissection identifies risk variants and genes related to artery integrity and tissue-mediated coagulation. Nat. Genet. 55, 964–972 (2023).

63. Pérez-Gutiérrez, L. & Ferrara, N. Biology and therapeutic targeting of vascular endothelial growth factor A. Nat. Rev. Mol. Cell Biol. 24, 816–834 (2023).

64. Velagapudi, S. et al. VEGF-A Regulates Cellular Localization of SR-BI as Well as Transendothelial Transport of HDL but Not LDL. Arterioscler. Thromb. Vasc. Biol. 37, 794–803 (2017).

65. Chen, H. X. & Cleck, J. N. Adverse effects of anticancer agents that target the VEGF pathway. Nat. Rev. Clin. Oncol. 6, 465–477 (2009).

66. Tall, A. R., Thomas, D. G., Gonzalez-Cabodevilla, A. G. & Goldberg, I. J. Addressing dyslipidemic risk beyond LDL-cholesterol. J. Clin. Invest. 132, e148559 (2022).

67. Zanoni, P. et al. Rare variant in scavenger receptor BI raises HDL cholesterol and increases risk of coronary heart disease. Science 351, 1166–1171 (2016).

68. Kurki, M. I. et al. FinnGen provides genetic insights from a well-phenotyped isolated population. Nature 613, 508–518 (2023).

69. Ritchie, S. C. et al. The Biomarker GlycA Is Associated with Chronic Inflammation and Predicts Long-Term Risk of Severe Infection. Cell Syst. 1, 293–301 (2015).

70. Lotta, L. A. et al. Genetic Predisposition to an Impaired Metabolism of the Branched-Chain Amino Acids and Risk of Type 2 Diabetes: A Mendelian Randomisation Analysis. PLoS Med. 13, e1002179 (2016).

71. Zhao, Y. et al. Small rodent models of atherosclerosis. Biomed. Pharmacother. Biomedecine Pharmacother. 129, 110426 (2020).

72. Shim, J., Al-Mashhadi, R. H., Sørensen, C. B. & Bentzon, J. F. Large animal models of atherosclerosis--new tools for persistent problems in cardiovascular medicine. J. Pathol. 238, 257–266 (2016).

73. Watanabe, K. et al. A global overview of pleiotropy and genetic architecture in complex traits. Nat. Genet. 51, 1339–1348 (2019).

74. Smith, C. J. et al. Integrative analysis of metabolite GWAS illuminates the molecular basis of pleiotropy and genetic correlation. eLife 11, e79348 (2022).

75. Julkunen, H. et al. Atlas of plasma NMR biomarkers for health and disease in 118,461 individuals from the UK Biobank. Nat. Commun. 14, 604 (2023).

76. Bycroft, C. et al. The UK Biobank resource with deep phenotyping and genomic data. Nature 562, 203–209 (2018).

77. Sudlow, C. et al. UK biobank: an open access resource for identifying the causes of a wide range of complex diseases of middle and old age. PLoS Med. 12, e1001779 (2015).

78. Karczewski, K. J. et al. Pan-UK Biobank GWAS improves discovery, analysis of genetic architecture, and resolution into ancestry-enriched effects. medRxiv (2024) doi:10.1101/2024.03.13.24303864.

79. Würtz, P. et al. Quantitative Serum Nuclear Magnetic Resonance Metabolomics in Large-Scale Epidemiology: A Primer on -Omic Technologies. Am. J. Epidemiol. 186, 1084–1096 (2017).

80. Ritchie, S. C. et al. Quality control and removal of technical variation of NMR metabolic biomarker data in ∼120,000 UK Biobank participants. Sci. Data 10, 64 (2023).

81. Buuren, S. van & Groothuis-Oudshoorn, K. mice: Multivariate Imputation by Chained Equations in R. J. Stat. Softw. 45, 1–67 (2011).

82. Mbatchou, J. et al. Computationally efficient whole-genome regression for quantitative and binary traits. Nat. Genet. 53, 1097–1103 (2021).

83. Willer, C. J., Li, Y. & Abecasis, G. R. METAL: fast and efficient meta-analysis of genomewide association scans. Bioinformatics 26, 2190–2191 (2010).

84. Bulik-Sullivan, B. K. et al. LD Score regression distinguishes confounding from polygenicity in genome-wide association studies. Nat. Genet. 47, 291–295 (2015).

85. Wang, G., Sarkar, A., Carbonetto, P. & Stephens, M. A simple new approach to variable selection in regression, with application to genetic fine mapping. J. R. Stat. Soc. Ser. B Stat. Methodol. 82, 1273–1300 (2020).

86. Wakefield, J. Bayes factors for genome-wide association studies: comparison with P-values. Genet. Epidemiol. 33, 79–86 (2009).

87. Rossen, J. et al. MultiSuSiE improves multi-ancestry fine-mapping in All of Us whole-genome sequencing data. medRxiv (2024) doi:10.1101/2024.05.13.24307291.

88. Chang, C. C. et al. Second-generation PLINK: rising to the challenge of larger and richer datasets. GigaScience 4, 7 (2015).

89. Keller, M. C. Gene × environment interaction studies have not properly controlled for potential confounders: the problem and the (simple) solution. Biol. Psychiatry 75, 18–24 (2014).

90. Kerimov, N. et al. A compendium of uniformly processed human gene expression and splicing quantitative trait loci. Nat. Genet. 53, 1290–1299 (2021).

91. Ochoa, D. et al. The next-generation Open Targets Platform: reimagined, redesigned, rebuilt. Nucleic Acids Res. 51, D1353–D1359 (2023).

92. Kanehisa, M. KEGG: Kyoto Encyclopedia of Genes and Genomes. Nucleic Acids Res. 28, 27–30 (2000).

93. Milacic, M. et al. The Reactome Pathway Knowledgebase 2024. Nucleic Acids Res. 52, D672–D678 (2024).

94. Murphy, A. E., Schilder, B. M. & Skene, N. G. MungeSumstats: a Bioconductor package for the standardization and quality control of many GWAS summary statistics. Bioinformatics 37, 4593–4596 (2021).

95. Wallace, C. Eliciting priors and relaxing the single causal variant assumption in colocalisation analyses. PLOS Genet. 16, e1008720 (2020).

96. Wallace, C. A more accurate method for colocalisation analysis allowing for multiple causal variants. PLOS Genet. 17, e1009440 (2021).

97. Uhlén, M. et al. Tissue-based map of the human proteome. Science 347, 1260419 (2015).

98. Giambartolomei, C. et al. Bayesian Test for Colocalisation between Pairs of Genetic Association Studies Using Summary Statistics. PLoS Genet. 10, e1004383 (2014).

99. Burgess, S., Small, D. S. & Thompson, S. G. A review of instrumental variable estimators for Mendelian randomization. Stat. Methods Med. Res. 26, 2333–2355 (2017).

100. Benjamini, Y. & Hochberg, Y. Controlling the False Discovery Rate: A Practical and Powerful Approach to Multiple Testing. J. R. Stat. Soc. Ser. B Stat. Methodol. 57, 289–300 (1995).

101. Backman, J. D. et al. Exome sequencing and analysis of 454,787 UK Biobank participants. Nature 5GG, 628–634 (2021).

102. McLaren, W. et al. The Ensembl Variant Effect Predictor. Genome Biol. 17, 122 (2016).

103. Ioannidis, N. M. et al. REVEL: An Ensemble Method for Predicting the Pathogenicity of Rare Missense Variants. Am. J. Hum. Genet. GG, 877–885 (2016).

104. Kircher, M. et al. A general framework for estimating the relative pathogenicity of human genetic variants. Nat. Genet. 46, 310–315 (2014).

105. Karczewski, K. J. et al. The mutational constraint spectrum quantified from variation in 141,456 humans. Nature 581, 434–443 (2020).

