## Supplemental figures 1-16 for "A genetic map of human metabolism across the allele frequency spectrum"

A

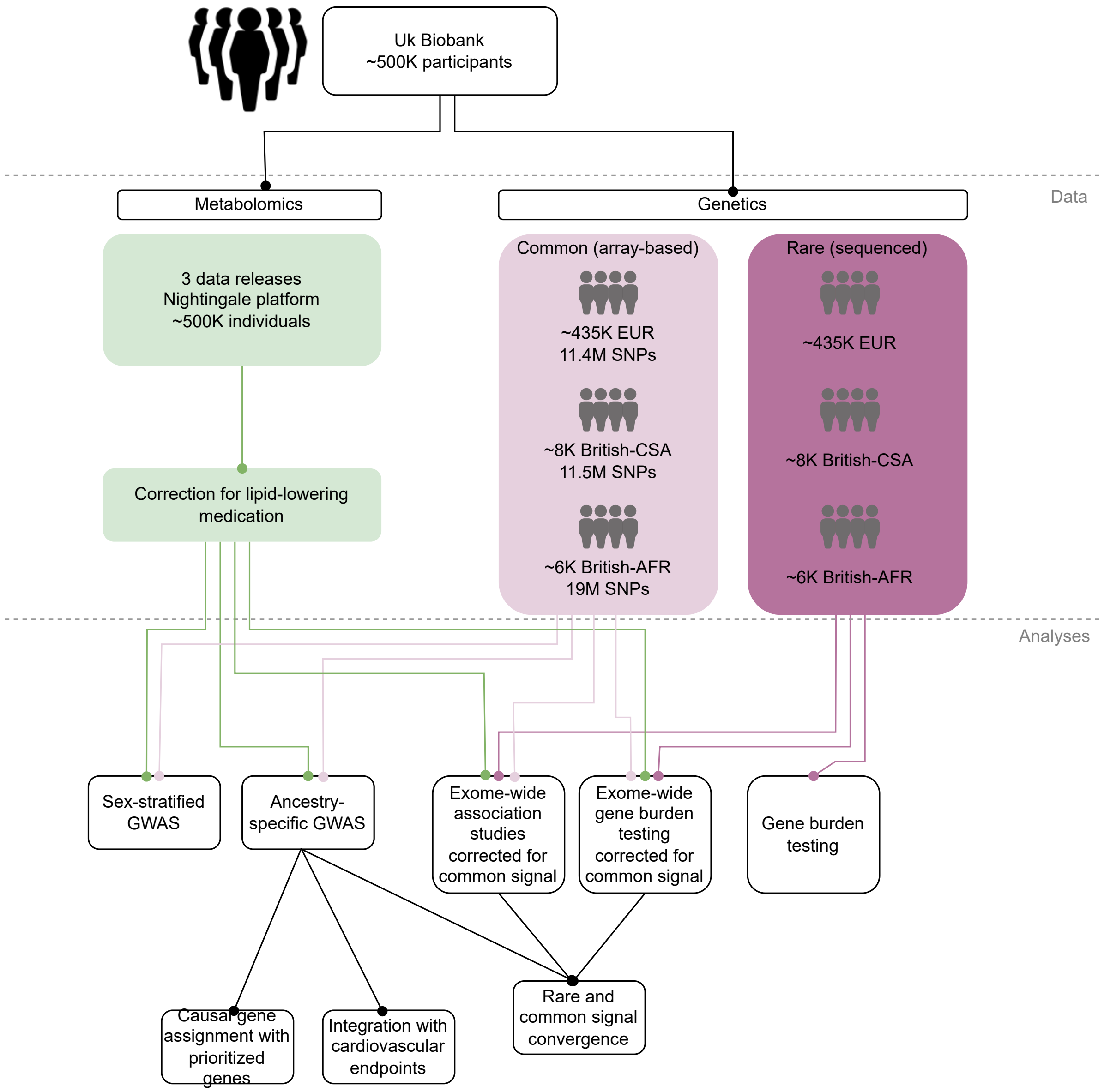


**Supplemental figure 1.** Graphical outline of the study design.

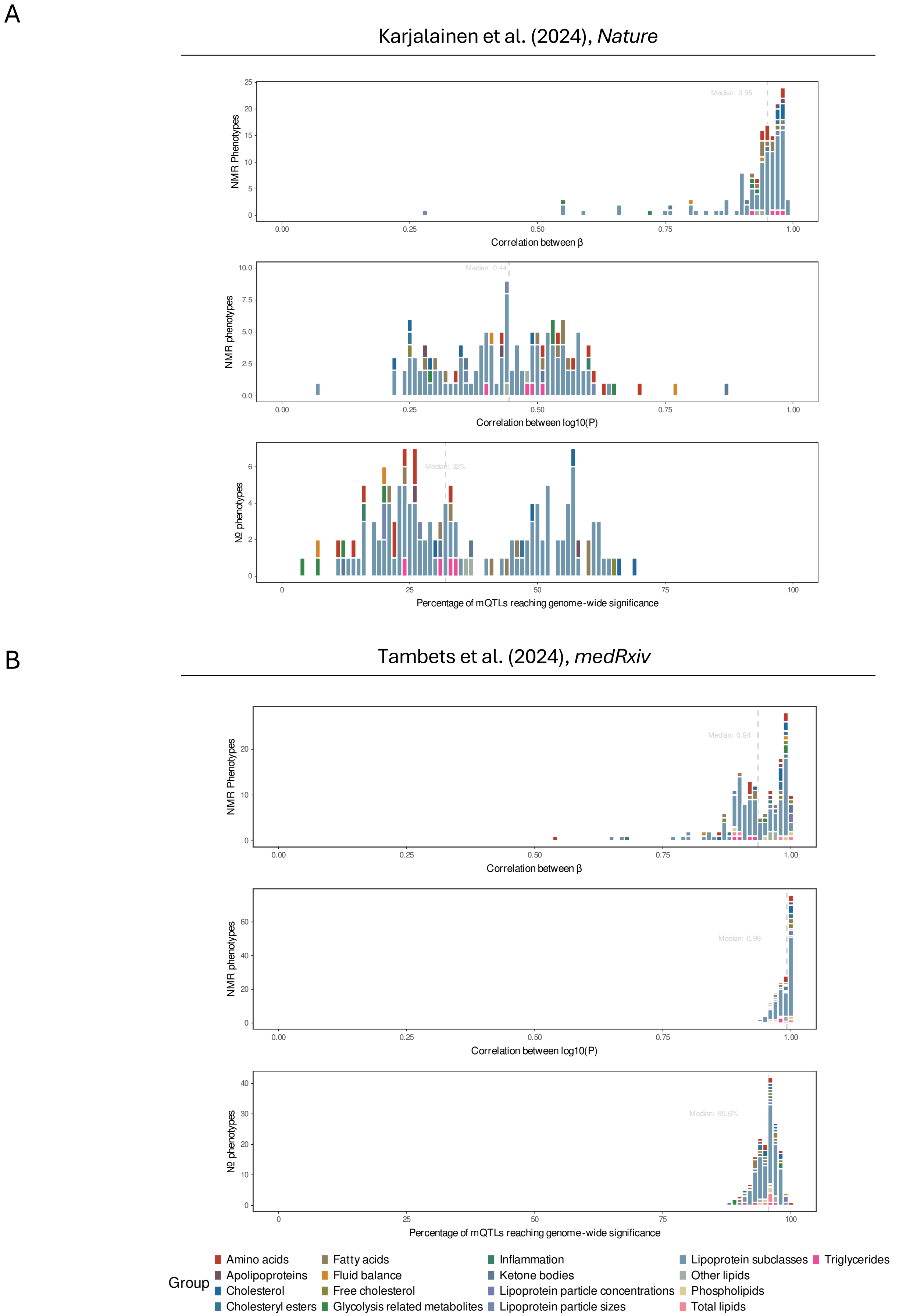


**Supplemental figure 2. A)** Replication of estimated genetic effects on circulating metabolites in Karjalainen et al., *Nature* (2024). Barplots represent the correlation of effect sizes (top), correlation between the P-value (middle), and the fraction of our sentinel variants that reached genome-wide significance in the replication study (bottom). **B)** Identical to **A)**, but using data from Tambets et al., *medRxiv* (2024). For both comparisons, we only considered directly measured traits.

A

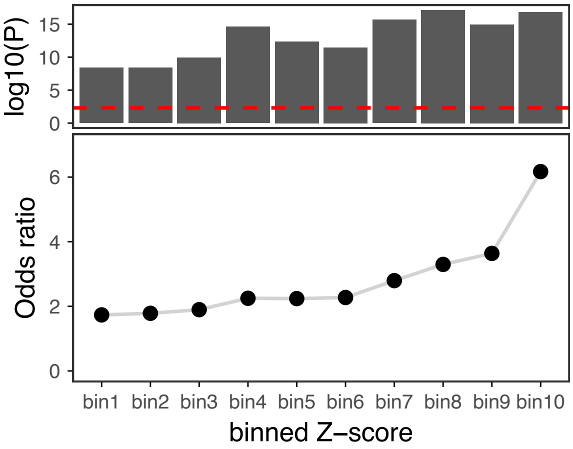


**Supplemental figure 3.** Enrichment of prioritised metabolic genes throughout bins of significance. Absolute Z-scores were binned in deciles and we extracted a list of genes proximal to fine-mapped lead variants. Enrichment of prioritised metabolic genes was calculated against the entire protein-coding background (n = 19747 genes)

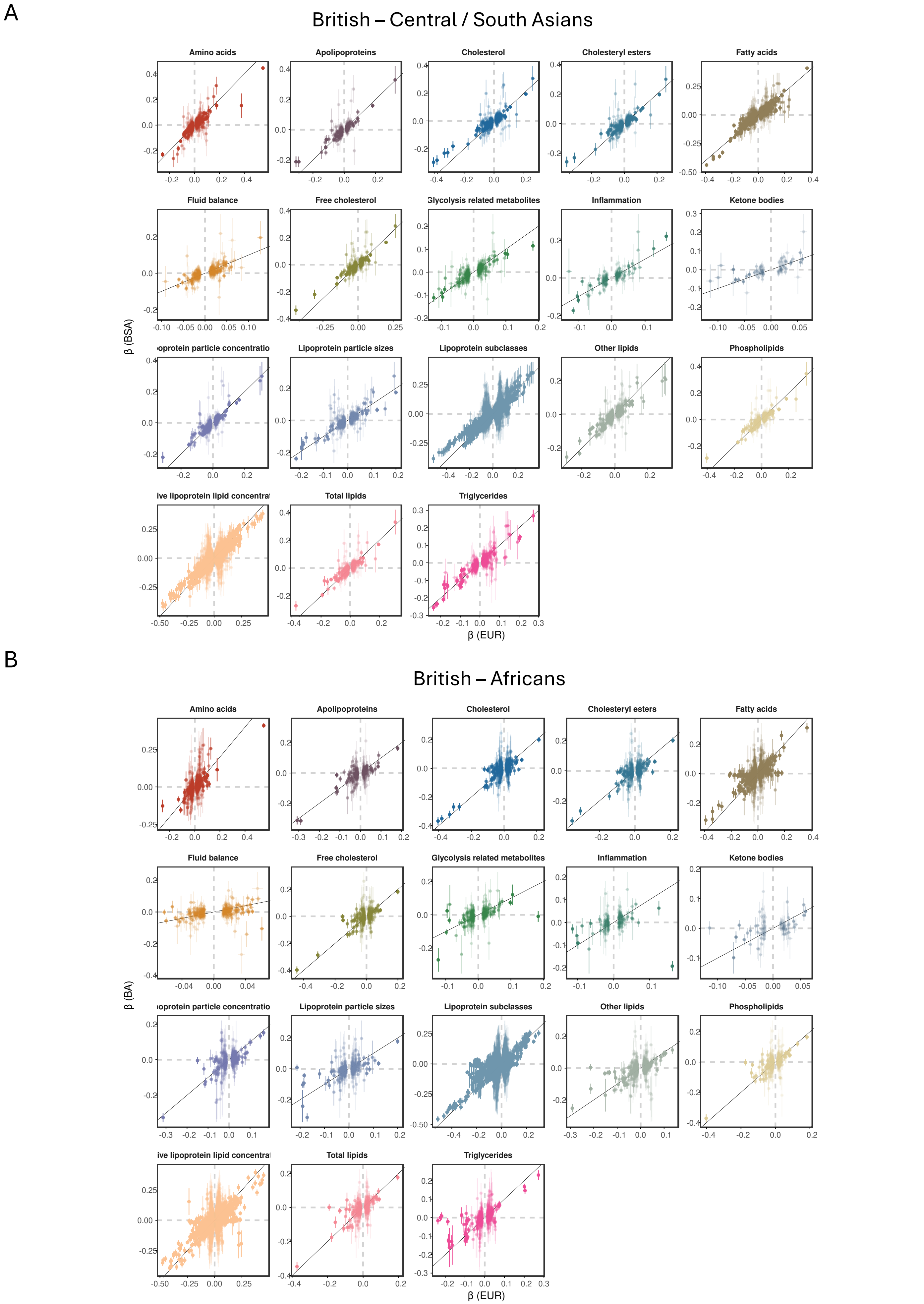


**Supplemental figure 4.** Cross-ancestry comparison of estimated genetic effects. Comparing estimates obtained in European participants (x-axis) to British-Asians (**A**) and British-Africans (**B**).

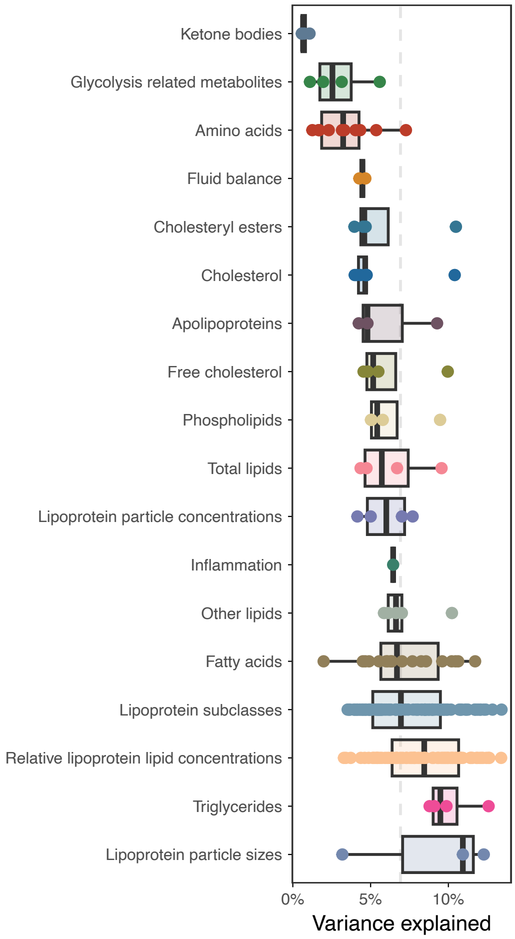


**Supplemental figure 5.** Variance explained by fine-mapped lead variants on metabolomic concentrations. Each dot represents a metabolite, colored for biochemical class.

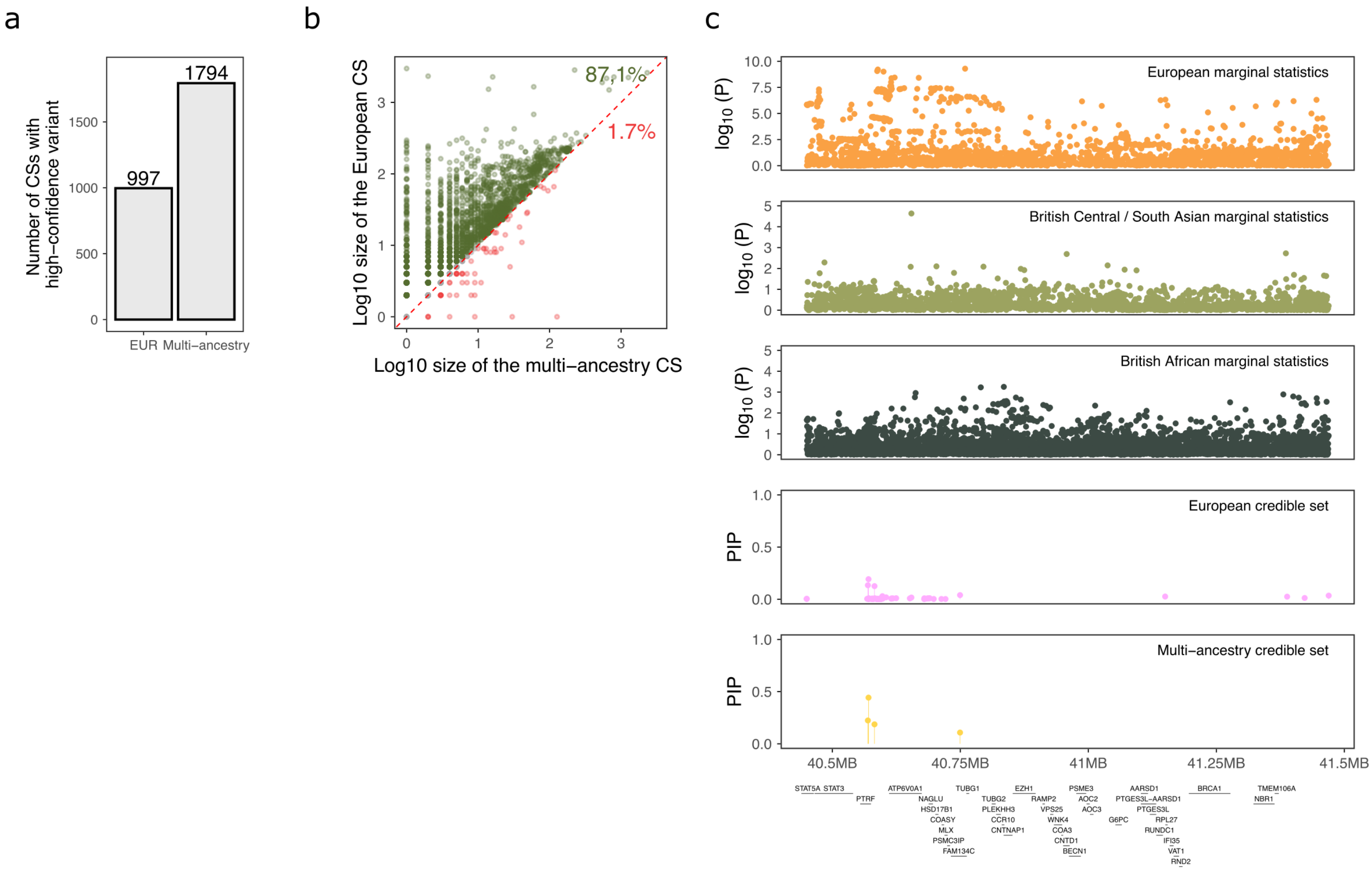


**Supplemental Figure 6: Refinement of credible sets using multi-ancestral genetic data. A)** Credible sets containing a high-confidence variant (posterior inclusion probability > 0.5) in the European-only and multi-ancestry fine-mapping analyses. **B)** Log-transformed size of European fine-mapped credible sets compared to multi-ancestral credible sets, shown for the credible sets that were considered in multi-ancestral finemapping analyses. The diagonal line represents equal credible set size between Europeans and multi-ancestral analyses, points above the diagonal indicate refined credible sets in multi-ancestral analyses. Percentages indicate the proportion of assessed credible sets that reduced in size through trans-ancestral finemapping (green) or increased (red). **C)** Example of a credible set before and after multi-ancestry fine-mapping, leading to assignment of a biologically plausible effector gene *PTRF*.

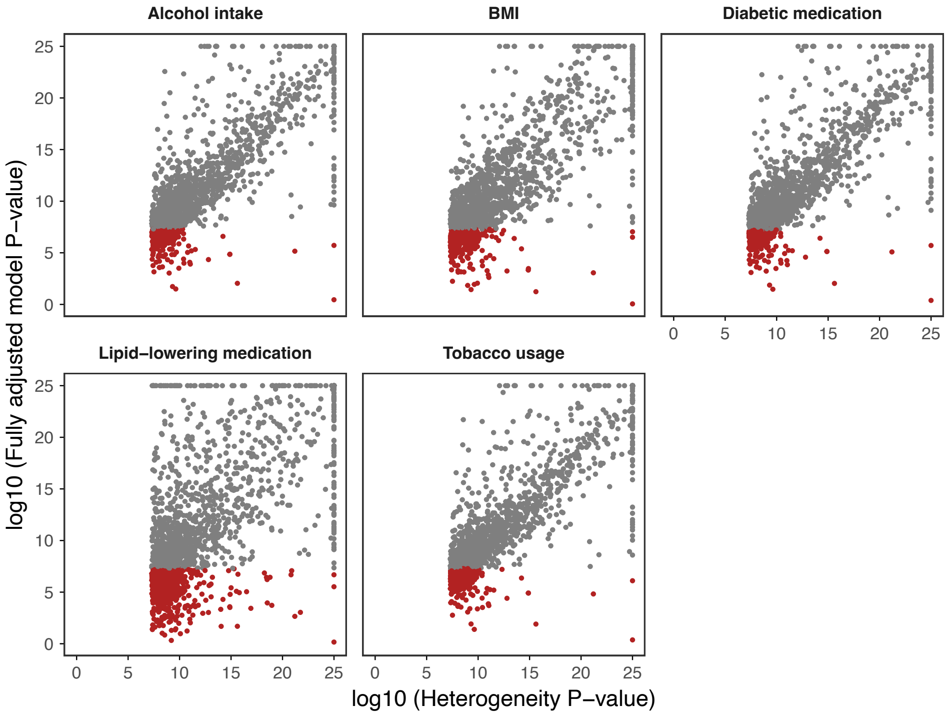


**Supplementary figure 7.** Comparison of P-values from 1800 putatively sex-differential loci identified through meta-analysis across the sexes. Shown are the METAL heterogeneity P-values (x-axis) and the p-values from the fully adjusted confounder model (y axis, see methods). Red dots represent loci where the sex-different effect was attenuated in the adjusted model and thus could be attributed to confounding effects.

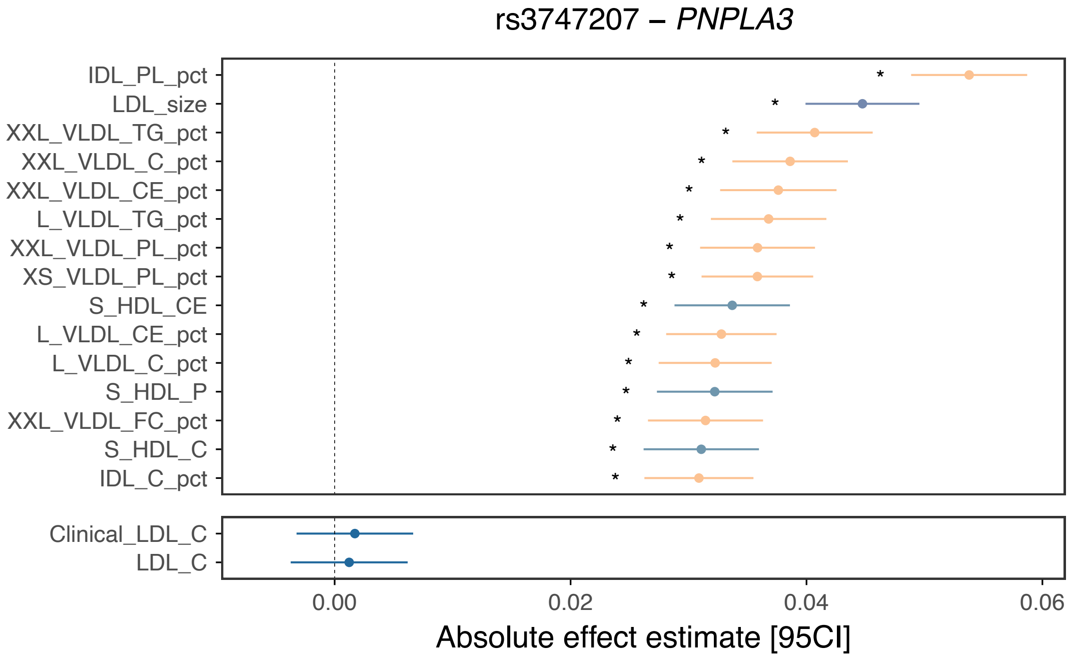


**Supplementary figure 8.** Forest plot showing the strongest associated NMR traits for rs3747207, previously associated to LDL-cholesterol. Stars represent whether traits are significantly differently associated compared to LDL-cholesterol.

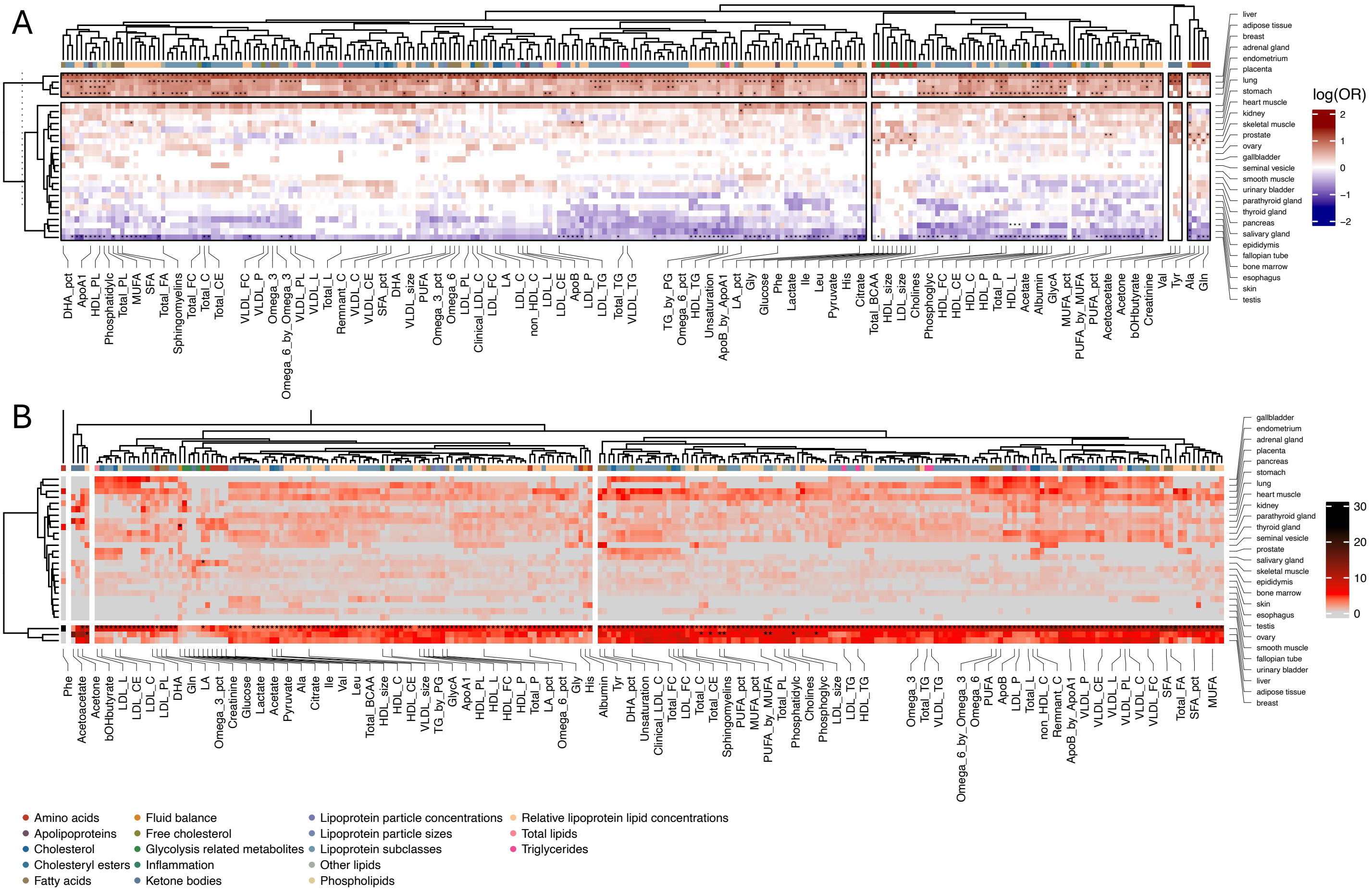


**Supplemental figure 9.** Odds ratios for enrichment of assigned effector genes **(A)** and genes proximal to fine-mapped lead variants **(B)** across tissue compartments. Columns represent each of the 249 metabolic traits, annotated for biochemical class. Rows and columns were clustered based on Euclidean distance.

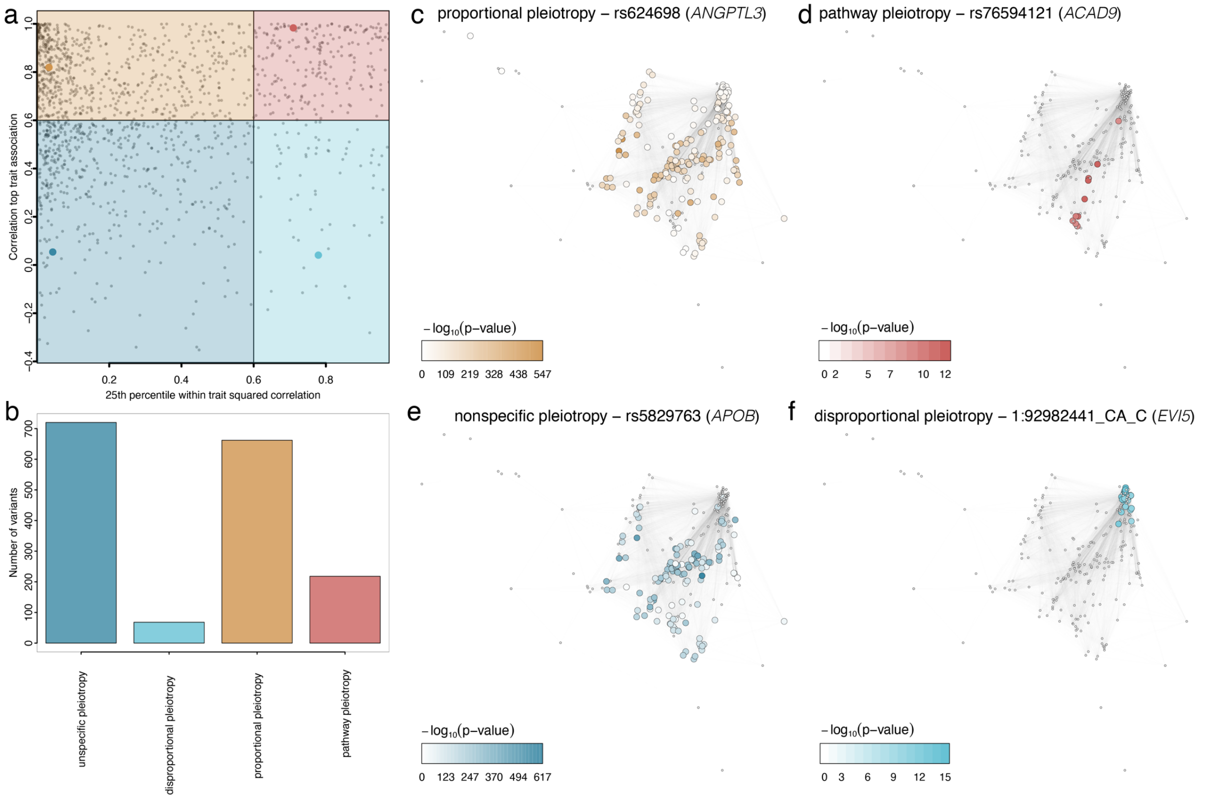


**Supplemental figure 10 a)** Scatterplot opposing mQTL characteristics. The x-axis denotes for each mQTL the 25$^{th}$ percentile of all possible correlations among associated NMR measures. The y-axis depicts the correlation between the strongest trait of interest and the association strength for all other traits. A value of one would indicate that all other associated NMR measures can be directly explained as function of correlation, whereas a value of zero would indicate independent effects of the mQTL on different measures. **c-f)** Same Pearson correlation networks of NMR measures, clustering highly correlated traits by spatial proximity. Each node is coloured according to the strength of associations (-log$_{10}$(p-value)) with one of the four genetic variants indicated in the title of each plot. Variants were chosen to represent each of the four modes of pleiotropy.

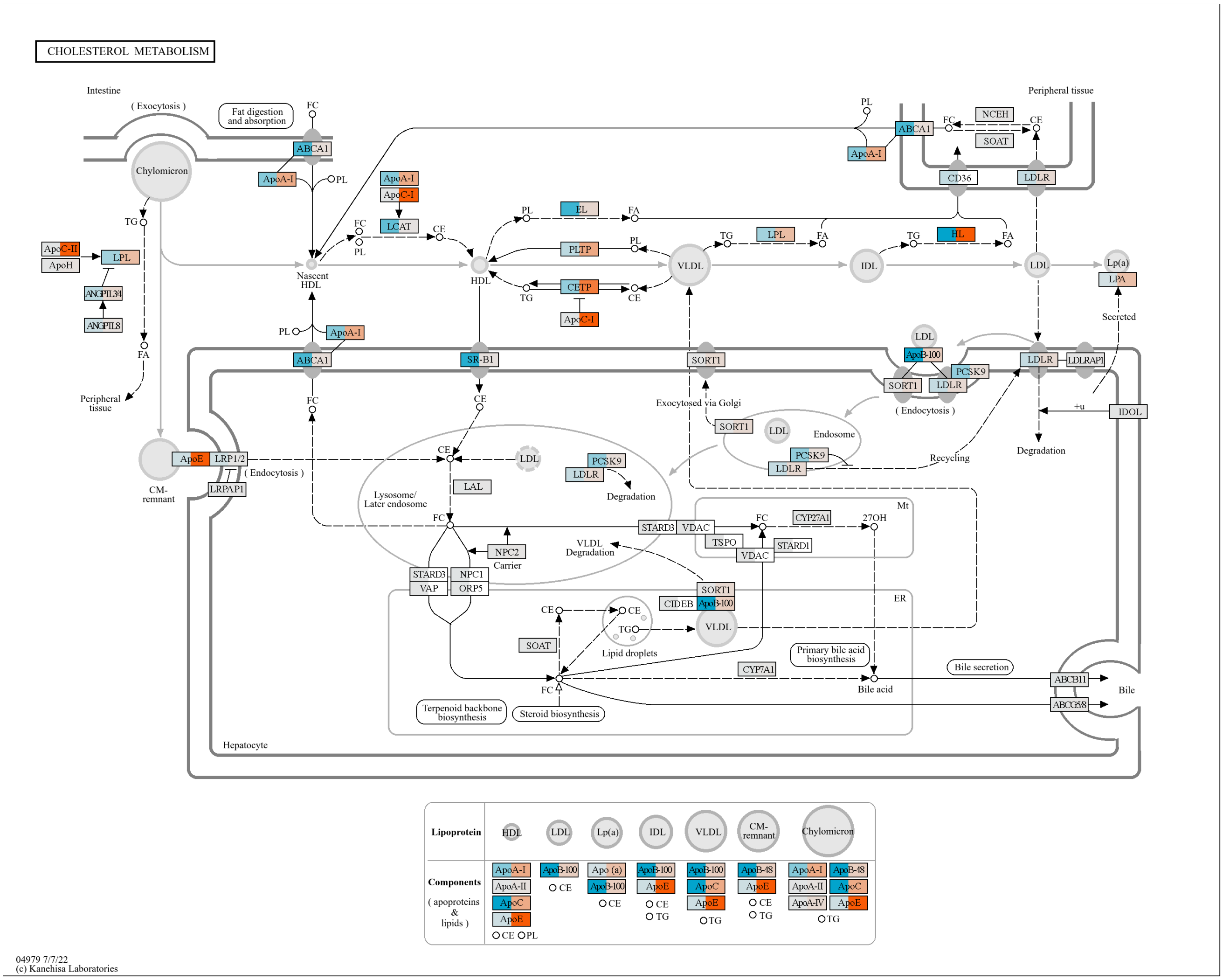


**Supplemental figure 11** Convergence of gene burden (blue) and common variant (orange) burden results for genes involved in cholesterol metabolism.

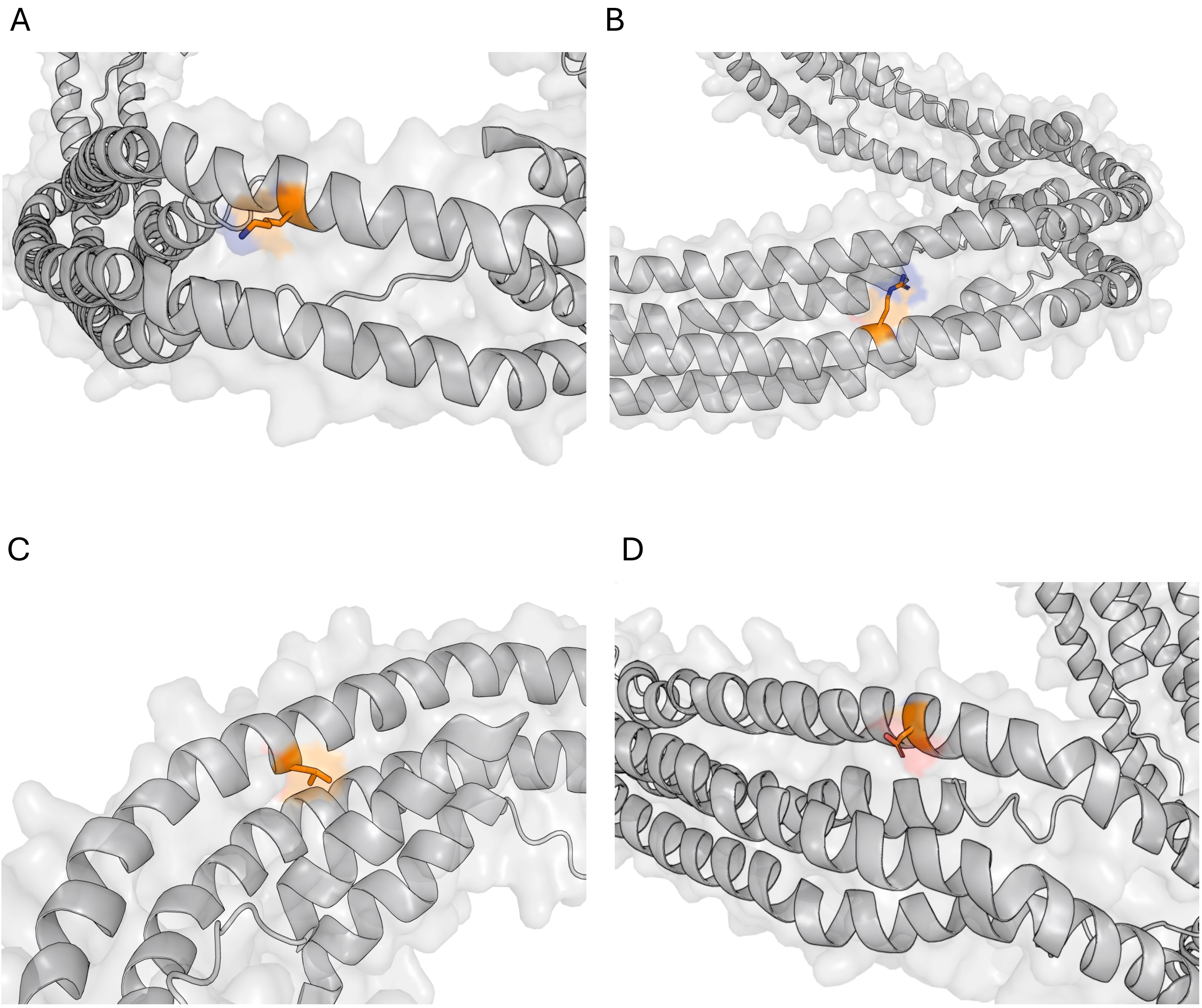


**Supplemental figure 12**. Location of 4 coding variants in APOA1 associated with NMR measures (PDB-ID 1AV1) coloured in orange; (**A**) p.Lys131del (**B**) p.Arg201Ser (**C**) p.Leu158Pro (**D**) p.Asp113Glu

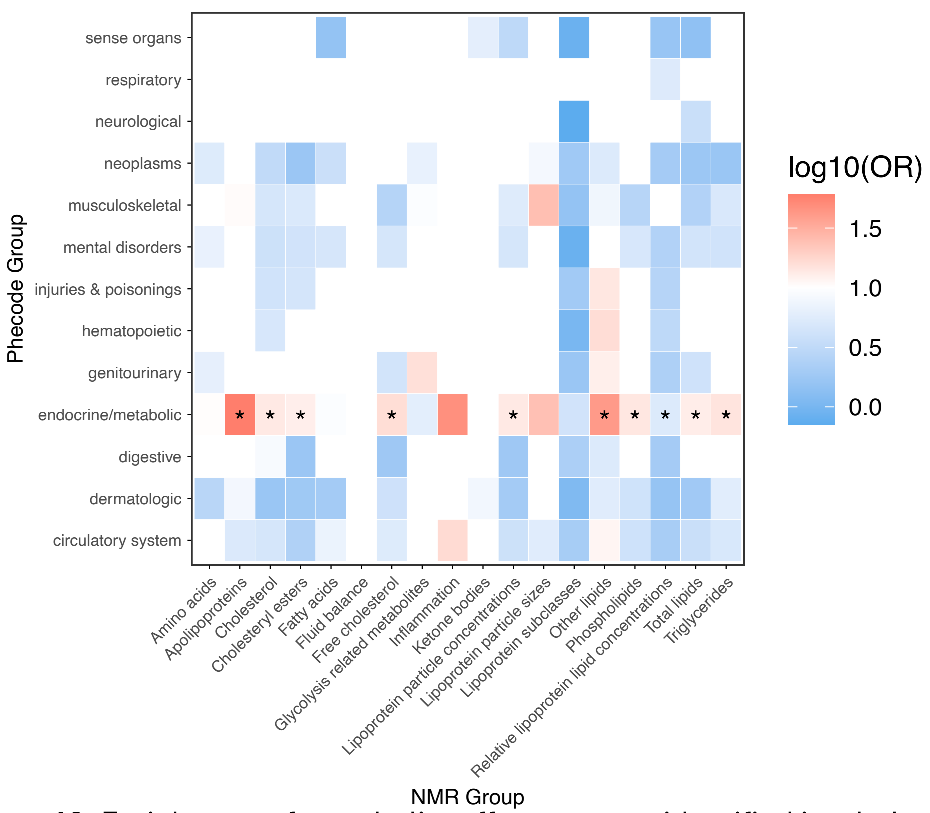


**Supplemental figure 13:** Enrichment of metabolite effector genes identified in whole-exome sequencing data amonst disease-related genes.

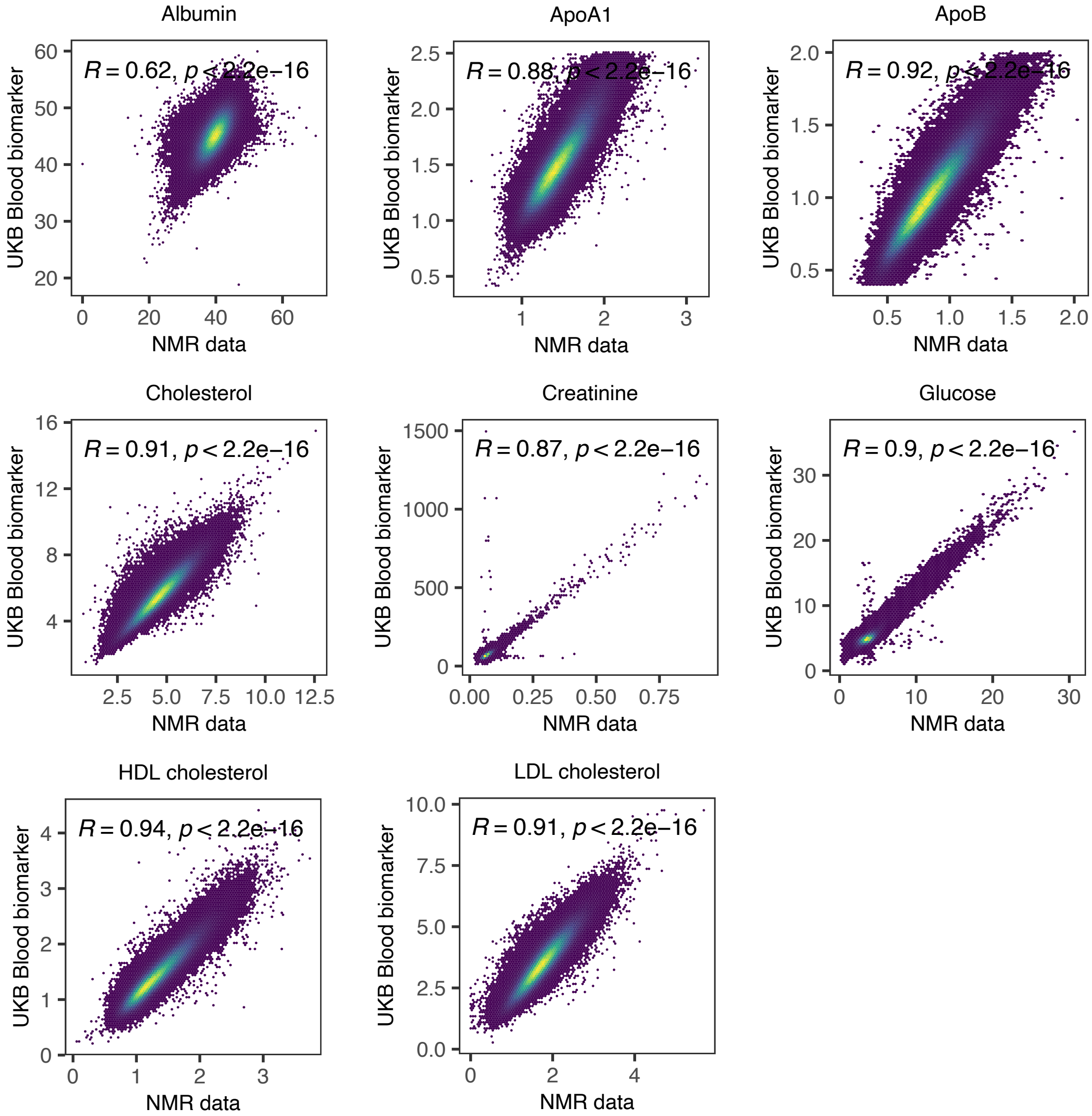


**Supplemental figure 14.** Comparison of 8 metabolic traits measured on the NMR platform (x axis) overlapping with routine blood biomarkers previously measured in the same cohort (y axis).

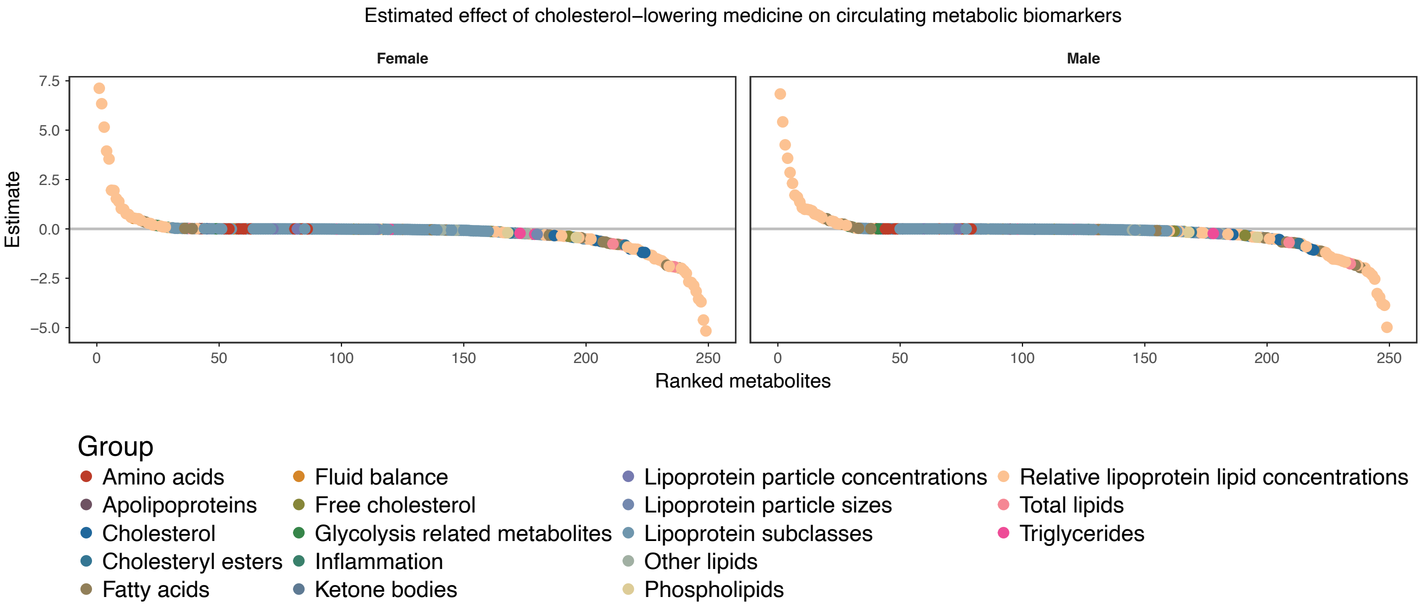


**Supplemental figure 15:** Effects of lipid-lowering medication on circulating metabolites. Each dot indicates one of 249 metabolites, ranked by the estimated effect of lipid-lowering medication on this metabolite. Metabolites are colored per biochemical class.

### Gene Assignment Classifier

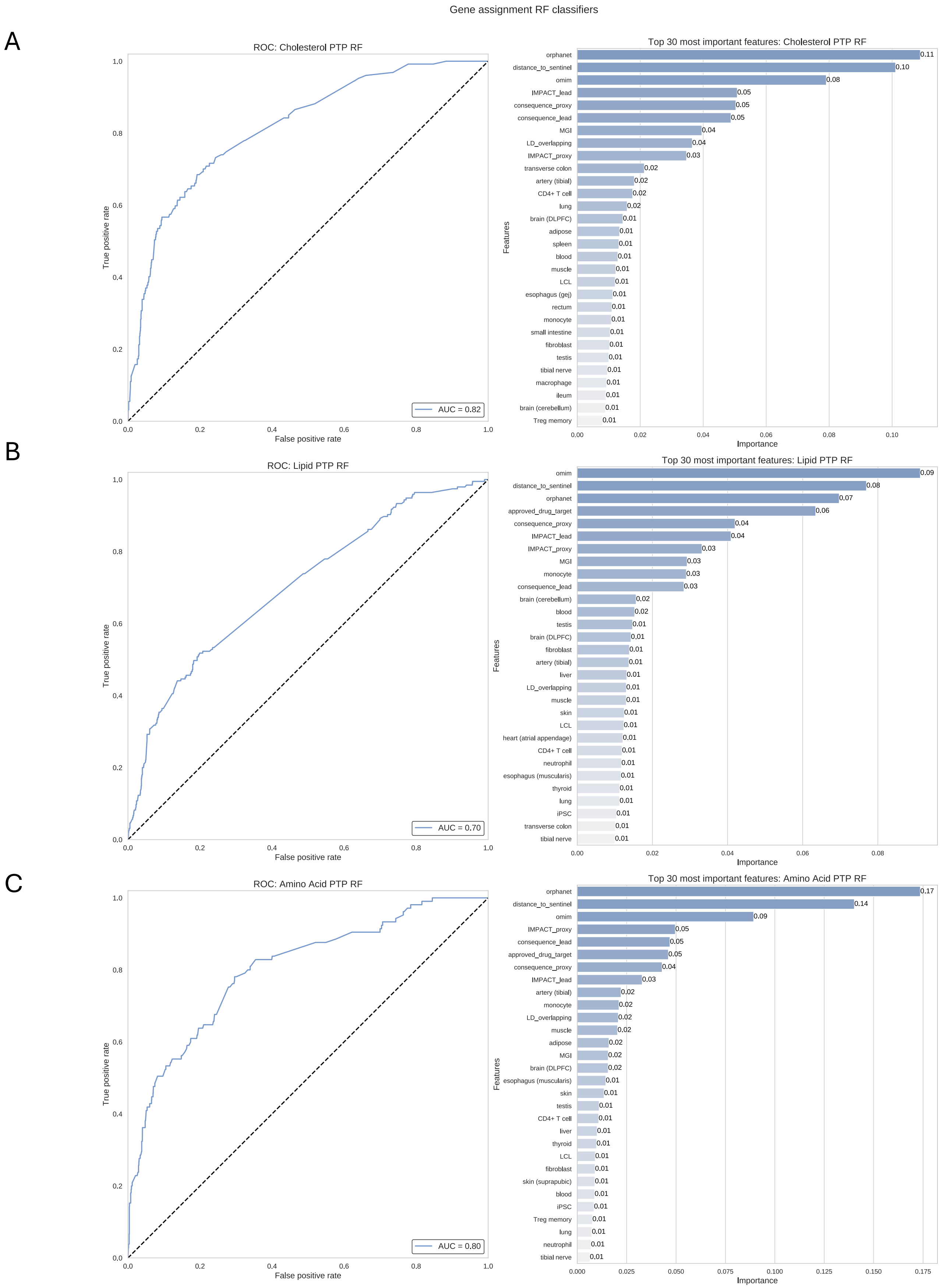


**Supplemental figure 16**. Area under the curve (left) and feature importance of the most predictive features (right) for models trained using a true positive set of genes annotated for cholesterol metabolism (**A**), lipid-related metabolism (**B**) and amino acid metabolism (**C**).
